# Safety and efficacy of long-term gantenerumab treatment in dominantly inherited Alzheimer’s disease: an open label extension of the phase 2/3 multicentre, randomised, double-blind, placebo-controlled platform DIAN-TU Trial

**DOI:** 10.1101/2024.10.29.24316289

**Authors:** Randall J. Bateman, Yan Li, Eric M. McDade, Jorge J. Llibre-Guerra, David B. Clifford, Alireza Atri, Susan L. Mills, Anna M. Santacruz, Guoqiao Wang, Charlene Supnet, Tammie L. S. Benzinger, Brian A. Gordon, Laura Ibanez, Gregory Klein, Monika Baudler, Rachelle S. Doody, Paul Delmar, Geoffrey A. Kerchner, Tobias Bittner, Jakub Wojtowicz, Azad Bonni, Paulo Fontoura, Carsten Hofmann, Luka Kulic, Jason Hassenstab, Andrew J. Aschenbrenner, Richard J. Perrin, Carlos Cruchaga, Alan E. Renton, Chengjie Xiong, Alison A. Goate, John C. Morris, David M. Holtzman, B. Joy Snider, Catherine Mummery, William S. Brooks, David Wallon, Sarah B. Berman, Erik Roberson, Colin L. Masters, Douglas R. Galasko, Suman Jayadev, Rachel Sanchez-Valle, Jeremie Pariente, Justin Kinsella, Christopher H. van Dyck, Serge Gauthier, Ging-Yuek Robin Hsiung, Mario Masellis, Bruno Dubois, Lawrence S. Honig, Clifford R. Jack, Alisha Daniels, David Aguillón, Ricardo Allegri, Jasmeer Chhatwal, Gregory Day, Nick Fox, Edward Huey, Takeshi Ikeuchi, Mathias Jucker, Jae-Hong Lee, Allan I. Levey, Johannes Levin, Francisco Lopera, JeeHoon Roh, Pedro Rosa-Neto, Peter R. Schofield, Dominantly Inherited Alzheimer’s Disease-Trials Unit

## Abstract

**Background:** Amyloid-plaque removal by monoclonal antibody therapies slows clinical progression in symptomatic Alzheimer’s disease; however, the potential for delaying the onset of clinical symptoms in asymptomatic people is unknown. The Dominantly Inherited Alzheimer Network Trials Unit (DIAN-TU) is an ongoing platform trial assessing the safety and efficacy of multiple investigational products in participants with dominantly inherited Alzheimer’s disease (DIAD) caused by mutations. On the basis of findings of amyloid removal and downstream biological effects from the gantenerumab arm of the platform trial, we continued a 3-year open-label extension (OLE) study to assess the safety and efficacy of long-term treatment with high doses of gantenerumab.

**Methods:** The randomised, placebo-controlled, double-blind, phase 2/3 multi-arm trial (DIAN-TU-001) assessed solanezumab or gantenerumab versus placebo in participants who were between 15 years before to 10 years after their estimated years to symptom onset and had a Clinical Dementia Rating (CDR) global score of 0 (cognitively normal) to 1 (mild dementia). This study was followed by an OLE study of gantenerumab treatment, conducted at 18 study sites in Australia, Canada, France, Ireland, Puerto Rico, Spain, the UK, and USA. For inclusion in the OLE, participants at risk for DIAD had participated in the double-blind period of DIAN-TU-001 and were required to know their mutation status. We investigated increasing doses of gantenerumab up to 1500 mg subcutaneous every 2 weeks. Due to the lack of a regulatory path for gantenerumab, the study was stopped early after a pre-specified interim analysis (when most participants had completed 2 years of treatment) of the clinical measure CDR-SB. The primary outcome for the final analysis was the amyloid plaque measure PiB-PET SUVR at 3 years, assessed in the modified intention to treat population (defined as participants who received any gantenerumab treatment post-OLE baseline, had at least one PiB-PET SUVR assessment prior to gantenerumab treatment, and a post-baseline assessment). All participants who received at least one dose of study drug in the OLE were included in the safety analysis. DIAN-TU-001 (NCT01760005) and the OLE (NCT06424236) are registered with clinicaltrials.gov.

**Findings:** Of 74 participants who were recruited into the OLE study between June 3, 2020 and April 22, 2021, 73 were enrolled and received gantenerumab treatment. 47 (64%) stopped dosing due to early termination of the study by the sponsor, and 13 (18%) prematurely discontinued the study for other reasons. The mITT population for the primary analysis comprised 55 participants. At the interim analysis, the hazard ratio for clinical decline of CDR-SB in asymptomatic mutation carriers was 0.79 (n=53, 95% CI 0.47 to 1.32) for participants who were treated with gantenerumab in either the double-blind or OLE period (Any Gant), and 0.53 (n=22, 0.27 to 1.03) for participants who were treated with gantenerumab the longest (Longest Gant). At the final analysis, the adjusted mean change from OLE baseline to year 3 in PiB-PET SUVR was -0.71 SUVR (95% CI -0.88 to -0.53, p<0.0001). Amyloid-related imaging abnormalities occurred in 53% (39/73) of participants: 47% (34/73) with microhaemorrhages, 30% (22/73) with oedema, and 6% (5/73) were associated with symptoms. No treatment-associated macrohaemorrhages or deaths occurred.

**Interpretation:** Partial or short-term amyloid removal did not show significant clinical effects. However, long-term full amyloid removal potentially delayed symptom onset and dementia progression. Conclusions are limited due to the OLE design and use of external controls and need to be confirmed in long term trials.

**Funding:** National Institutes on Aging, Alzheimer’s Association, GHR, F. Hoffmann-La Roche, Ltd/Genentech

## INTRODUCTION

Alzheimer’s disease is a devastating neurodegenerative disorder with a growing global health burden. Decades of research support the amyloid hypothesis, which posits that cerebral accumulation of amyloid β plaques is a key initiating event in the pathogenesis of Alzheimer’s disease.^1,2^ While monoclonal antibody therapies, including aducanumab, lecanemab, and donanemab, have been shown to reduce amyloid-plaque deposition and slow cognitive and functional decline at the group level in early symptomatic Alzheimer’s disease (mild cognitive impairment or mild Alzheimer’s disease dementia), their ability to prevent or delay the onset of cognitive impairment in at risk individuals is unknown.^3–5^

Dominantly inherited Alzheimer’s disease (DIAD) is caused by autosomal dominant mutations in presenilin-1 (*PSEN1*), presenilin-2 (*PSEN2*), or the amyloid-beta precursor protein (*APP*) genes that alter APP processing, resulting in overproduction of aggregation prone amyloid β and accumulation of amyloid β-plaque pathology in early to mid-adulthood.^6–8^ Carriers of these mutations have a predictable age of symptom onset and follow a preclinical stage characterised by amyloid β-plaque deposition decades before cognitive decline, providing a unique model for investigating disease-modifying therapies in a prevention paradigm.^9^

In 2012, the Dominantly Inherited Alzheimer Network Trials Unit (DIAN-TU) launched the first prevention trial (DIAN-TU-001, NCT01760005) to investigate the efficacy of two anti-amyloid monoclonal antibodies in parallel (solanezumab and gantenerumab) in delaying symptom onset and slowing progression in asymptomatic and mildly symptomatic DIAD mutation carriers. While the double-blind period of the DIAN-TU-001 phase 3 trial (2012–2019) with gantenerumab did not meet its primary endpoint of slowing cognitive decline in both symptomatic and asymptomatic cohorts, it significantly reduced amyloid-plaques in a dose-dependent fashion, reduced CSF total tau and tau phosphorylated at threonine 181 (p-tau181), and attenuated increases of CSF neurofilament light chain, a non-specific marker of neurodegeneration, particularly in asymptomatic DIAD mutation carriers.^10^ Further examination of downstream CSF and blood-based biomarkers showed that gantenerumab treatment significantly improved multiple CSF measures of synaptic degeneration, microglial activity, other inflammatory markers, and astrocyte activation markers.^11^ Together, these data provided the rationale to continue with a 3-year open-label extension study (OLE, 2020-2023, NCT06424236), with the goal of determining if continued treatment with gantenerumab at substantially higher doses than those used in sporadic Alzheimer’s disease studies and the DIAN-TU double-blind period could result in complete removal of brain amyloid plaques, improve critical markers of disease progression, improve or normalise non-amyloid related downstream biomarkers, and impact expected clinical and cognitive measures and trajectories.

The GRADUATE I and II trials of gantenerumab in symptomatic sporadic Alzheimer’s disease did not demonstrate significant slowing of clinical decline.^12^ This outcome could be attributed to the fact that the magnitude of amyloid plaque removal was smaller than predicted, with only one quarter of participants treated with gantenerumab reaching amyloid negative status at the end of the trial.^12^ As the DIAN-TU gantenerumab OLE was ongoing when the GRADUATE results were reported, and the cohort had been treated for approximately 8 years at that time, we established a pre-specified interim analysis to determine whether substantial clinical benefit of slowed disease progression and amyloid removal was achieved and whether continuation to complete the study was justified. Here we report the interim and final results from the DIAN–TU-001 gantenerumab OLE trial for the pre-specified primary and secondary outcomes.

## METHODS

### Study design

The DIAN-TU-001 was a randomised, double-blind, placebo-controlled, phase 2/3 trial of solanezumab or gantenerumab versus placebo, with a common close design, conducted from December 2012 to November 2019.^10^ Briefly, the DIAN-TU-001 was a 2-year biomarker study that was transitioned to a 4-year treatment trial with a cognitive primary end point to investigate the potential for amyloid-removing drugs to slow or prevent cognitive decline. This gantenerumab OLE was designed to continue to treat eligible participants from the DIAN-TU-001 double-blind period with substantially increasing high doses of gantenerumab for 3 years. There was a mean treatment gap of approximately 1 year (0.4 SD) between the end of the double-blind period and the start of the OLE period in 2020 (Figure 1). Recruitment took place between June 3, 2020 and April 22, 2021, at 18 DIAN-TU study sites in Australia, Canada, France, Ireland, Puerto Rico, Spain, UK, and continental United States of America (US). The study included a pre-specified interim analysis involving selected clinical, cognitive, and biomarker endpoints to determine: (i) if high dose or long term, or both, gantenerumab treatment resulted in clinical benefit, and (ii) whether the high doses of gantenerumab resulted in substantial amyloid removal compared to the double-blind period. The interim analysis was conducted when most participants had received 2 years of high dose gantenerumab treatment (data lock March 31, 2023). Due to the lack of a conclusive major clinical benefit and regulatory path for gantenerumab, dosing was stopped in August 2023, and the study was closed by the sponsor.

**Figure 1.**
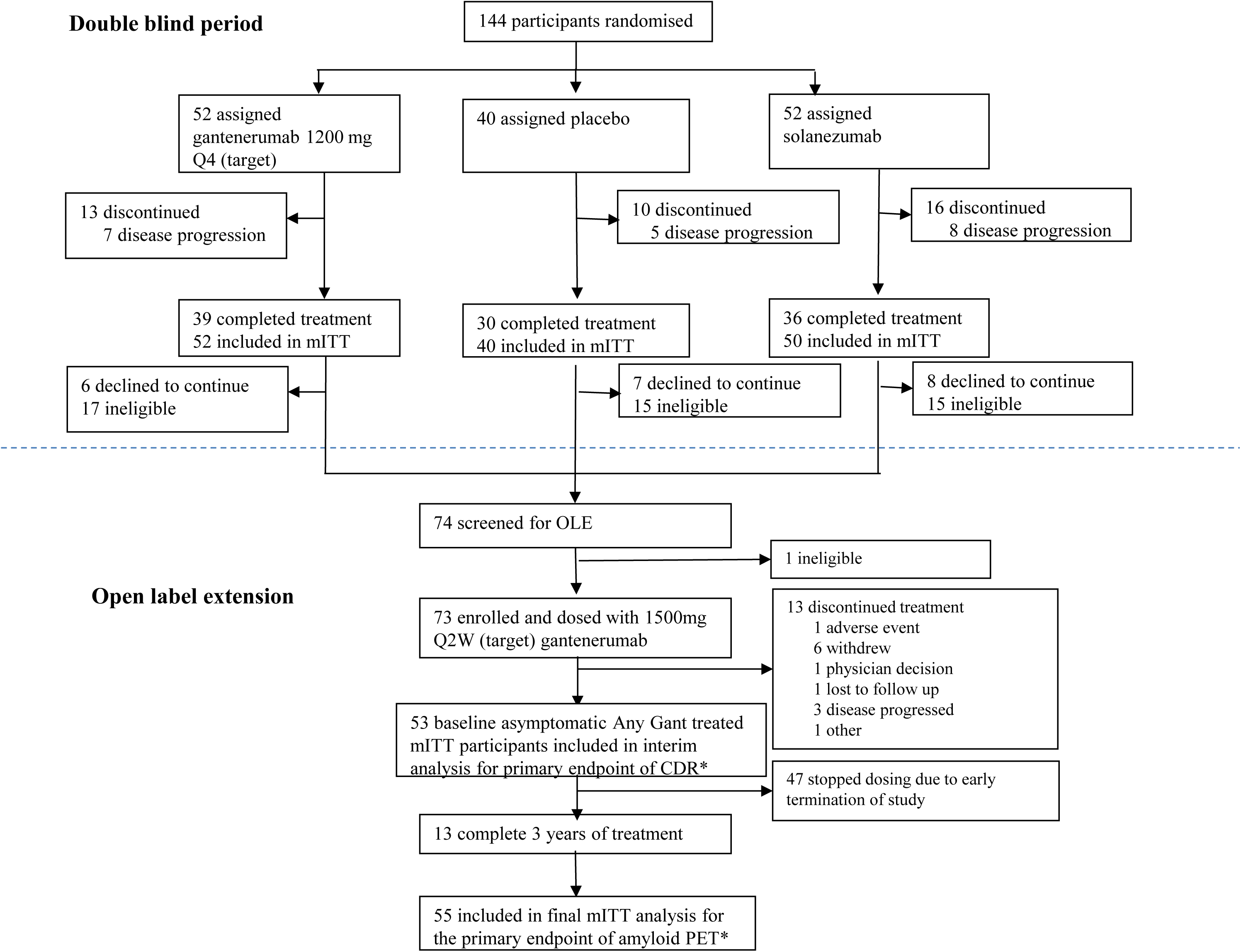
Trial profile. Gantenerumab/solanezumab double-blind and gantenerumab open-label extension arms of the DIAN-TU-001 prevention trial. The sample sizes presented are for the mITT population of the primary endpoint for interim analysis (n=53) and final analysis (n=55). Participants who met inclusion criteria and were analysed according to the pre-specified primary analyses are shown. See Supplemental Figure 7 for participant retention, drop-out, and duration of treatment. *All participants did not have all outcome measurements, thus the sample size is different for different endpoints.

Ethics approval was obtained through a central Institutional Review Board (Advarra) for US sites (main approved protocol number Pro0003095). Ethics committee approval was obtained at all other participating sites.

### Participants

For original enrolment in the DIAN-TU double-blind period, participants were between 15 years before to 10 years after (inclusive) of their estimated years to symptom onset (EYO = participant’s age at the clinical assessment – participant’s mutation and parental estimated age at symptom onset),^13^ and had a Clinical Dementia Rating (CDR) global score of 0 (cognitively normal) to 1 (mild dementia) (Figure 1). Participant mutation status and *APOE* genotype were determined using Clinical Laboratory Improvement Amendments (CLIA) whole exome sequencing.

For enrolment in the OLE, participants had to have participated in the double-blind period and were willing to know their mutation status. The main exclusion criteria included major or unstable illness that would prevent trial participation or completion of main study-related testing, volumetric MRI contraindications, required anticoagulation therapy, or pregnancy. Other reasons for exclusion included advanced disease, death, or early termination from the double-blind period for other reasons. A small number were unwilling to know their mutation status and declined participation. Participants provided written informed consent.

Participant data with a cutoff date of June 30, 2022 from the DIAN Observational study (DIAN Obs, NCT00869817)^9,14^ and that met the DIAN-TU-001 trial inclusion criteria were used as natural history external controls for improved estimates of the placebo group in the primary analyses. Internal controls are the data from placebo treated participants from the double-blind phase who did not enroll in the OLE period. Of note, the DIAN Obs and DIAN-TU studies are harmonised, including use of the same clinical, cognitive, imaging, and biomarker measures. Participant groups for analysis included the main set of controls, defined as the external controls (DIAN Obs) plus the internal controls (placebo treated in the double-blind phase) (Supplemental Figure 5A-C). The extended set controls (dashed box on Supplemental Figure 5B) are defined as the main set controls plus the double-blind baseline and OLE baseline values for participants treated with placebo during the double-blind period and who entered the OLE.

### Randomisation and masking

Randomisation and group assignments for the DIAN-TU-001 double-blind phase have been published.^10^ Briefly, mutation carriers were assigned 3:3:2 to receive gantenerumab (n=52), solanezumab (n=52), or placebo (n=40). Randomisation was done with a minimisation procedure by central randomisation. Allocation concealment was performed through central randomization. For the gantenerumab OLE, participants were enrolled at DIAN-TU study sites by study coordinators and individuals involved in the conduct of the study who were kept masked to the participants’ treatment assignments in the double-blind period, and were not involved in data analyses.

### Procedures

During the double-blind period, participants received increasing doses of gantenerumab (Generation 3 drug product), from 225 mg and up to 1200 mg subcutaneous (SC) administration every 4 weeks, or placebo. During the OLE phase, gantenerumab (Generation 4 drug product used in the GRADUATE trials) doses were titrated from 120mg every 4 weeks to up to 1020 mg or 1500 mg every 2 weeks (see Supplemental Figure 1 for dose titration schedule). The 1020 mg Generation 4 dose was PK-equivalent of the 1200 mg Generation 3 dose. The average titration time to the highest doses was 19 months (SD 3.6), with 57 of 73 (78%) participants completing titration to the highest dose. The planned duration of treatment in the OLE was 3 years. To reduce participant burden, home health nurses were used for most drug administration visits. The average treatment compliance rate is 93%.

To determine if gantenerumab at high doses would result in the removal of brain amyloid, participants underwent PiB-PET at OLE baseline and annually with CSF and blood collections, clinical assessments with CDR, and cognitive assessments with MMSE and cognitive batteries. Details of the assessments are presented in Supplemental Methods 2.

### Outcomes

The pre-specified primary endpoints for the interim analysis were the time to recurrent progression in CDR-SB and time to first progression in CDR global. The CDR scale stages dementia severity by assessing six domains: Memory, Orientation, Judgment and Problem Solving, Community Affairs, Home and Hobbies, and Personal Care. A CDR global score of 0 represents normal or no cognitive impairment, 0.5 very mild, 1 mild, 2 moderate, and 3 severe dementia. Time to first progression in CDR global is defined as the time from OLE baseline to the first visit where CDR increases. Progression must be observed at two consecutive measurements unless progression occurs at the final measurement. CDR-SB is an 18-point ordinal measure of dementia severity, with higher scores representing greater severity. Recurrent progression of CDR-SB is any progression which is either the first increase in value above baseline or any progression above the highest preceding post-baseline value identified as a progression, where progression above the preceding value must be observed at two consecutive measurements unless the progression occurs at the last measurement. For example, if a participant’s CDR-SB is 0, 0.5, 0.5 and 1 at baseline, year 1, 2 and 3, respectively, then this participant had two progressions: 0 to 0.5 from baseline to year 1, and 0.5 to 1 from year 2 to 3. Event time is defined as time since baseline for the first progression and time to previous event for subsequent progressions. Participants without a progression event at their last measurement were censored at the time of that measurement. Other outcomes used for interim analysis are detailed in Supplemental Methods 4.

The primary outcome measure for the final analysis was the change in amount of amyloid-plaque deposition as measured by the PiB-PET composite. The pre-specified key secondary outcome was CDR global and CDR-SB progression as defined at the interim analysis.

Other secondary endpoints were: (i) DIAN-TU Cognitive Composite (Supplemental Method 3), with lower scores interpreted as worse performance; (ii) Functional Assessment Scale (FAS), which measures the participant’s ability to perform daily activities, with higher scores indicating worse performance; (iii) MMSE with lower scores indicate worse performance. For FAS and MMSE, increases of 3 units or more was considered a progression and time to recurrent progression was defined similarly as CDR-SB. (iv) Biomarkers CSF Aβ 1-42/1-40, with lower values indicating abnormality; and Tau-PET SUVR and CSF p-tau181/tau181, with higher values indicating abnormality. (v) CSF neurofilament light chain (NfL) analysis is in progress and will be reported once available.

ARIA was a safety endpoint in this study. Surveillance MRI was built into the dose escalation design to monitor for ARIA events routinely, and any suspected ARIA was assessed by the medical monitoring team promptly (see Supplemental Figure 1). Other safety outcomes included the incidence and severity of treatment-emergent adverse events (TEAEs), serious adverse events (SAEs), and treatment discontinuations. Clinical laboratory evaluations, vital signs, and 12-lead electrocardiograms were also measured throughout the study.

These endpoints were developed and finalised by the DIAN-TU, Roche, and the U.S. Food and Drug Administration (FDA), and pre-specified in the Statistical Analysis Plan.

### Statistical Analysis

For the final analysis, the primary analysis was performed for the OLE modified intent-to-treat (mITT) population, defined as participants who received any gantenerumab treatment post-OLE baseline, had at least one PiB-PET SUVR assessment prior to gantenerumab treatment, and a post-baseline assessment (based on OLE period data only, referred to as “OLE only Gant”, Supplementary Figure 5C). We estimated annual dropout rates of 5% and 10% and anticipated approximately 56 to 43 participants to remain at the end of the 3-year OLE, and the study was predicted to have over 99% power to detect the projected mean change from baseline using a one-sided one-sample t-test with a type I error rate of 2.5% (see Supplemental Methods 2). The primary analysis of the primary endpoint of PiB-PET SUVR reduction from baseline in the OLE mITT population was performed using a mixed model for repeated measures (MMRM) analysis without controls. The MMRM included the OLE baseline PiB-PET SUVR and visit (treated as categorical) as fixed effects. An unstructured variance-covariance matrix was used to model the within-subject errors among the repeated measures. To demonstrate that gantenerumab significantly removes amyloid plaque at year 3, the upper bound of the 95% confidence interval at the year 3 OLE visit would be less than 0. To evaluate the treatment effect of gantenerumab during the whole study duration, the change from double-blind baseline to the end of the OLE period in PiB-PET SUVR by the double-blind treatment group was also evaluated using a similar MMRM model with fixed effects of double-blind baseline PiB-PET SUVR, visit, double-blind treatment group and its interaction with visit. The proportion of participants who converted from abnormal (PiB-PET SUVR > 1.25) to normal (PiB-PET SUVR ≤ 1.25) was also examined. The DIAN normal cutoff was 1.25 PiB-PET SUVR derived based on the 95^th^ percentile of PiB-PET SUVR from asymptomatic non-carriers.

The primary analysis for the key secondary endpoint of time to recurrent progression in CDR-SB, involved comparing participants who were treated with gantenerumab in either the double-blind or OLE period (referred to as “Any Gant”, Supplementary Figure 5A) to the main set controls. For all the comparisons using the Cox model stated below, start time was the baseline for the corresponding population as defined in Supplementary Figure 5A-C, end time was the last available CDR assessment time. To compare the probability of recurrent progression in CDR-SB between treated and controls, the Cox proportional hazards model was applied employing the Andersen-Gill method,^15^ a robust variance estimator to account for the correlation between recurrent events. The Cox model incorporated the group variable and baseline EYO as fixed effects in the analysis. Additionally, the Cox model was used to analyse the time to first progression in CDR Global. The Cox proportional hazards model generates hazard ratios that provide an estimate of the relative hazard of progression between treatment and control groups. The same Cox proportional hazard models were also applied to the comparison (i) between the longest gantenerumab mITT Population (“Longest Gant”, Supplementary Figure 5B) and the main set of controls, (ii) between Longest Gant mITT Population and the extended set of controls, (iii) between OLE only Gant mITT Population and the main set of control. Post-hoc sensitivity analyses based on CDR are described in Supplemental Method 1. As post-hoc analysis, similar Cox models were also applied to the FAS and MMSE to estimate the hazard ratios between Longest Gant mITT Population and the extended set of control.

MMRM models similar to those used for PiB-PET SUVR were employed to analyse the key secondary endpoints CSF p-tau181/tau181 and CSF Aβ 1-42/1-40. The Linear Mixed Effects (LME) model with random intercept and random slope was utilised to analyse other key secondary endpoints including the Cognitive Composite, FAS, MMSE and Tau-PET SUVR. For the LME analysis, the annual rate of change between the OLE mITT population and the main set controls (internal controls only for Tau-PET SUVR) was compared. The LME model incorporated time since baseline (treated as a continuous variable), treatment group, the interaction between treatment group and time as fixed effects, and random intercepts and slopes for each participant as random effects. Unstructured covariance matrix was used for random effects. CSF biomarkers were also converted to CentiMarker values utilising a similar methodology as the Centiloid conversion approach. For CentiMarker calculations, more abnormal biomarker raw values indicate worse disease stages as defined by the direction of disease versus normal groups. The detailed steps for calculating CentiMarker are described in Supplemental Method 6.

We conducted a pre-specified interim analysis with the data collected up to March 31, 2023, when most participants had completed 2 years of treatment in OLE. For the interim analysis, the same MMRM and Cox models as described for the final analysis were used for amyloid PET SUVR, CDR SB and global (see Supplementary Method 4 for details).

All analyses were conducted using SAS software, version 9.4.

The safety analysis population includes all participants who received at least one dose of study drug in the gantenerumab OLE period. A Data Safety Monitoring Board (DSMB) reviewed unblinded safety monitoring data throughout the trial, assessed risk-benefit ratio acceptability, provided periodic recommendations regarding the progress of the study, and reported to the central Institutional Review Board and participant site Ethics Committees. The OLE is registered in ClinicalTrials.gov, NCT06424236.

Other methods descriptions, including assessments, imaging and fluid biomarker methods are in Supplemental Methods 2.

## RESULTS

### Participant characteristics

Of the 144 mutation carriers enrolled in the DIAN-TU-001 double-blind study, 74 (52%) were successfully recruited into the gantenerumab OLE study between June 3, 2020 and April 22, 2021; 21 (15%) declined participation and 47 (33%) were ineligible to participate (Figure 1). One participant screen failed due to inability to receive drug and appropriate clinical safety assessments. The remaining 73 participants received gantenerumab treatment per protocol. Overall, 13 (18%) of these 73 participants completed the 3 years of treatment for the study; 47 (64%) stopped dosing due to early termination of the study by sponsor, and 13 (18%) prematurely discontinued the study for other reasons (Figure 1). The primary analysis population for PiB-PET SUVR included 55 mITT participants in gantenerumab OLE. The median duration of the OLE period was 2.64 years (IQR 1.99 to 2.90 years). For CDR analysis based on baseline asymptomatic participants (double-blind baseline for prior gantenerumab treated group and OLE baseline for prior placebo and solanezumab treated group), the Any Gant mITT population (Supplementary Figure 5A) included 53 participants, the Longest Gant treated mITT population (Supplementary Figure 5B) included 22 participants, the OLE only Gant treated mITT population included 40 participants (Supplementary Figure 5C), the main set of control group included 74 participants, and the extended control group (Supplementary Figure 5B) that included the OLE baseline measures, comprised 86 participants. Demographics and baseline characteristics at OLE or double-blind baseline are presented in Table 1. Length of follow up for different groups are presented in Supplemental Table 1.

**Table 1.** Demographics and baseline characteristics of the analysis populations for the gantenerumab open-label extension.

|  | Overall |  | Baseline Asymptomatic |  |  |  |  |
| --- | --- | --- | --- | --- | --- | --- | --- |
|  | OLE only Gant baseline<br>(n = 62) | Main set of control<br>(n = 144) | Any Gant treated mITT<br>(n = 53) | Longest Gant treated mITT<br>(n = 22) | OLE only Gant baseline<br>(n = 40) | Main set of control<br>(n = 74) | Extended set of control<br>(n = 86) |
| **Age** (years)<br>(mean ± SD) | 48.3 ± 8.03 | 41.7 ± 9.89 | 43.0 ± 8.07 | 43.1 ± 7.32 | 46.1 ± 7.20 | 37.24 ± 9.47 | 37.6 ± 9.11 |
| **Female** (n (%)) | 35 (56) | 81 (56) | 28 (53) | 11 (50) | 24 (60) | 46 (62) | 55 (64) |
| **Male** (n (%)) | 27 (44) | 63 (44) | 25 (87) | 11 (50) | 16 (40) | 28 (38) | 31 (36) |
| **Race** (n (%)) |  |  |  |  |  |  |  |
| White | 56 (90) | 112 (78) | 49 (92) | 21 (95) | 37 (93) | 55 (74) | 64 (74) |
| Black or African American | 0 | 1 (1) | 0 | 0 | 0 | 1 (1) | 1 (1) |
| Other | 6 (10) | 31 (22) | 4 (8) | 1 (5) | 3 (8) | 18 (24) | 21 (24) |
| **Ethnicity** (n (%)) |  |  |  |  |  |  |  |
| Hispanic | 8 (13) | 24 (17) | 8 (15) | 3 (14) | 7 (18) | 14 (19) | 17 (20) |
| Not Hispanic | 54 (87) | 119 (83) | 45 (85) | 19 (86) | 33 (83) | 59 (80) | 68 (79) |
| Unknown | 0 | 1 (1) | 0 | 0 | 0 | 1 (1) | 1 (1) |
| **Gene** (n (%)) |  |  |  |  |  |  |  |
| APP | 7 (11) | 22 (15) | 3 (6) | 0 (0) | 3 (7) | 9 (12) | 10 (12) |
| PSEN1 | 50 (81) | 113 (78) | 45 (85) | 19 (86) | 32 (80) | 58 (78) | 68 (79) |
| PSEN2 | 5 (8) | 9 (6) | 5 (9) | 3 (14) | 5 (13) | 7 (9) | 8 (9) |
| **APOE ε4** (n (%)) | 17 (27) | 43 (30) | 17 (32) | 4 (18) | 10 (25) | 18 (24) | 21 (24) |
| **EYO**<br>median (Q1, Q3) | 0.33 (-3.62, 4.01) | -2.91 (-11.71, 2.14) | -5.86 (-9.03, -0.34) | -7.16 (-10.53, -5.14) | -2.84 (-5.71, 0.58) | -10.77 (-13.59, -4.17) | -9.61 (-13.39, -4.39) |
| CDR 0 (n (%)) | 40 (65) | 74 (51) | 53 (100) | 22 (100) | 40 (100) | 74 (100) | 86 (100) |
| CDR 0.5 (n (%)) | 10 (16) | 54 (38) | 0 | 0 | 0 | 0 | 0 |
| CDR 1 (n (%)) | 10 (16) | 16 (11) | 0 | 0 | 0 | 0 | 0 |
| **CDR-SB**<br>median (Q1, Q3) | 0 (0, 2.00) | 0.50 (0, 2.50) | 0 (0, 0) | 0 (0, 0) | 0 (0, 0) | 0 (0, 0) | 0 (0, 0) |
| **FAS** median (Q1, Q3) | 0 (0, 1.11) | 0 (0, 3.75) | 0 (0, 0) | 0 (0, 0) | 0 (0, 0) | 0 (0, 0) | 0 (0, 0) |
| **Cognitive composite** | 0.45 (-0.13, 0.99) | 0.07 (-0.49, 0.61) | 0.67 (0.14, 1.04) | 0.58 (0.14, 1.02) | 0.78 (0.32, 1.12) | 0.48 (0.15, 0.86) | 0.50 (0.18, 0.85) |
| median (Q1, Q3) |  |  |  |  |  |  |  |
| **MMSE**<br>median (Q1, Q3) | 29.0 (27.0, 30.0) | 28.5 (25.0, 30.0) | 30.0 (28.0, 30.0) | 30.0 (29.0, 30.0) | 30.0 (29.0, 30.0) | 29.0 (29.0, 30.0) | 29.0 (29.0, 30.0) |
| **PiB-PET SUVR**<br>median (Q1, Q3) | 2.3 (1.6, 3.4) | 2.2 (1.5, 3.2) | 2.0 (1.2, 2.7) | 1.7 (1.5, 2.2) | 1.9 (1.3, 2.6) | 1.7 (1.1, 2.5) | 1.7 (1.2, 2.5) |
| **PiB-PET Centiloid**<br>median (Q1, Q3) | 48 (19, 96) | 52 (14, 90) | 44 (10, 65) | 26 (14, 48) | 31 (10, 63) | 26 (2, 59) | 32 (7, 59) |
| **Total dose (mg)**<br>median (Q1, Q3) | 61.4K (39.5K, 101.4K) | 0 | 62.4K (44.8K, 102.6K) | 110.5K (90.3K, 122.2K) | 68.6K (53.2K, 110.5K) | 0 | 0 |

<sup>a</sup>For Any Gant treated mITT, double-blind baseline was used for prior gantenerumab treated participants and OLE baseline was used for prior placebo and solanezumab treated participants. For Longest Gant treated mITT, double-blind baseline was used. For OLE only Gant, OLE baseline was used. Any Gant=participants treated with gantenerumab in either the double-blind or OLE period, *APOE* ε4=presence of at least one ε4 allele of apolipoprotein E, *APP*=amyloid precursor protein, CDR=Clinical Dementia Rating, CDR-SB=Clinical Dementia Rating – Sum of Boxes, EYO=estimated years to symptom onset, FAS=Functional Assessment Scale, Longest Gant=participants treated with gantenerumab in both the double-blind and OLE periods, mITT=modified intent-to-treat population, OLE only Gant=participants treated with gantenerumab in the OLE period, PiB-PET=<sup>11</sup>C-Pittsburgh compound-B positron emission tomography, *PSEN1*=presenilin-1, *PSEN2*=presenilin-2, SUVR=standardized uptake value ratio.
Sex was determined by participant self-report.

### Interim Analysis primary and secondary outcomes: clinical dementia progression and amyloid plaque removal

The primary outcomes for asymptomatic participants who received Any Gant (n = 53) had a hazard ratio (95% CI) of 0.91 (0.43, 1.94) for time to first progression in CDR global, and 0.79 (0.47, 1.32) for time to recurrent progression in CDR-SB compared to the main set control. For the Longest Gant treated baseline asymptomatic group (n = 22) with average duration of 8 years, the hazard ratio (95% CI) was 0.43 (0.15, 1.26) for CDR global and 0.53 (0.27, 1.03) for CDR-SB. For the OLE period, the OLE only Gant baseline asymptomatic group with average duration of 2 years (n=40), the hazard ratio (95% CI) was 1.39 (0.46, 4.19) for CDR global and 1.41 (0.59, 3.33) for CDR-SB (see Supplemental Table 2). Amyloid-plaque removal, measured by PiB-PET SUVR, was dose-dependent, with approximately 3-fold greater rates of amyloid-plaque lowering by tripling the dose used in the double-blind period (Supplemental Figure 2). However, only 8% (4/49) of participants converted from amyloid abnormal to normal and 12% (6/49) of participants stayed amyloid normal (defined by SUVR<1.25).

### Final Analysis Primary efficacy endpoint: amyloid plaque removal

The adjusted mean change from OLE baseline (mean 2.62, SD 1.30) to year 3 (mean 1.96, SD 1.12) was PiB-PET SUVR of -0.71 (95% CI -0.88 to -0.53, p < 0.0001, Supplemental Figure 3, shown in Centiloids in Figure 2A). Significant reductions were also observed in PiB-PET SUVR from OLE baseline to the end of OLE year 1 (-0.12, 95% CI -0.21 to -0.03, p = 0.012) and OLE year 2 (-0.47, 95% CI -0.60 to -0.33, p<0.0001) (Supplemental Figure 3). Participants who received higher doses of gantenerumab (1020-1500 mg every 2 weeks) showed approximately 3-fold greater rates of amyloid plaque lowering, with annual reduction of 0.35 SUVR (OLE year 1 to 2) compared to those on lower doses (225 mg every 4 weeks), with annual reduction of 0.06 SUVR (Supplemental Figure 2). At the end of the OLE, 8 (15%) participants remained amyloid PET normal (defined by SUVR<1.25)^16^, 8 (15%) participants converted from abnormal to normal, and 38 (70%) participants remained amyloid PET abnormal (Supplemental Table 8). In comparison, the group that was treated with gantenerumab the longest achieved 45% amyloid plaque negative (normal) status, with 55% remaining amyloid PET abnormal.

**Figure 2.**
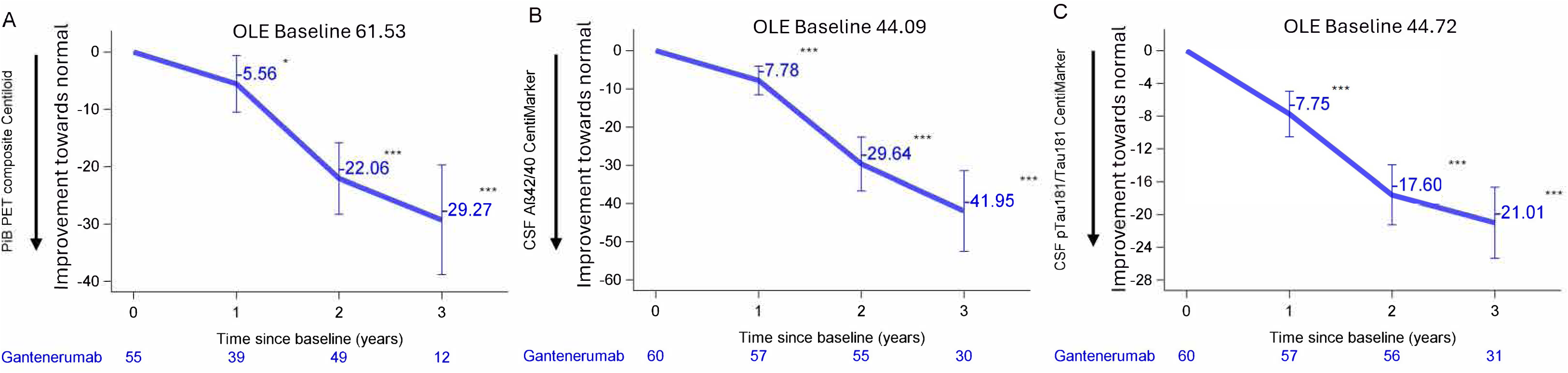
Primary and secondary biomarker outcomes. Estimated mean change from open-label extension (OLE) baseline (95% confidence intervals) for the treatment group using mixed model for repeated measures analyses for (A) the primary outcome of the OLE - brain amyloid burden measured by the average standardized uptake value ratio (SUVR) of cortical regions of interest (superior frontal, rostral middle frontal, superior temporal, middle temporal, lateral orbitofrontal, medial orbitofrontal and precuneus), assessed by ^11^C-Pittsburgh compound-B positron emission tomography (PiB-PET) and shown in Centiloids, (B) cerebrospinal fluid (CSF) Aβ42/40, and (C) CSF phospho-tau181/non-phosphorylated tau. \**P* < 0.05, \*\**P* < 0.01, \*\*\**P* < 0.001.

At OLE year 3, a significant reduction in mean SUVR relative to the double-blind baseline was demonstrated for OLE participants from the gantenerumab treated arm (-0.44, 95% CI -0.76 to - 0.12) and solanezumab treated arm (-0.47, 95% CI -0.86, -0.09), but not for OLE participants from the placebo arm (-0.07, 95% CI -0.51, 0.36) (Supplemental Figure 3).

### Final Analysis Secondary outcomes: key clinical dementia progression

In 53 participants who were asymptomatic at baseline and were included in Any Gant, the hazard ratio (95% CI) of time to first progression in CDR global score for treatment group vs. the main set of control was 0.84 (0.40 to 1.77), and the hazard ratio for time to recurrent progression in CDR-SB was 0.83 (0.50 to 1.36) (Figure 3, Supplemental Table 3).

**Figure 3.**
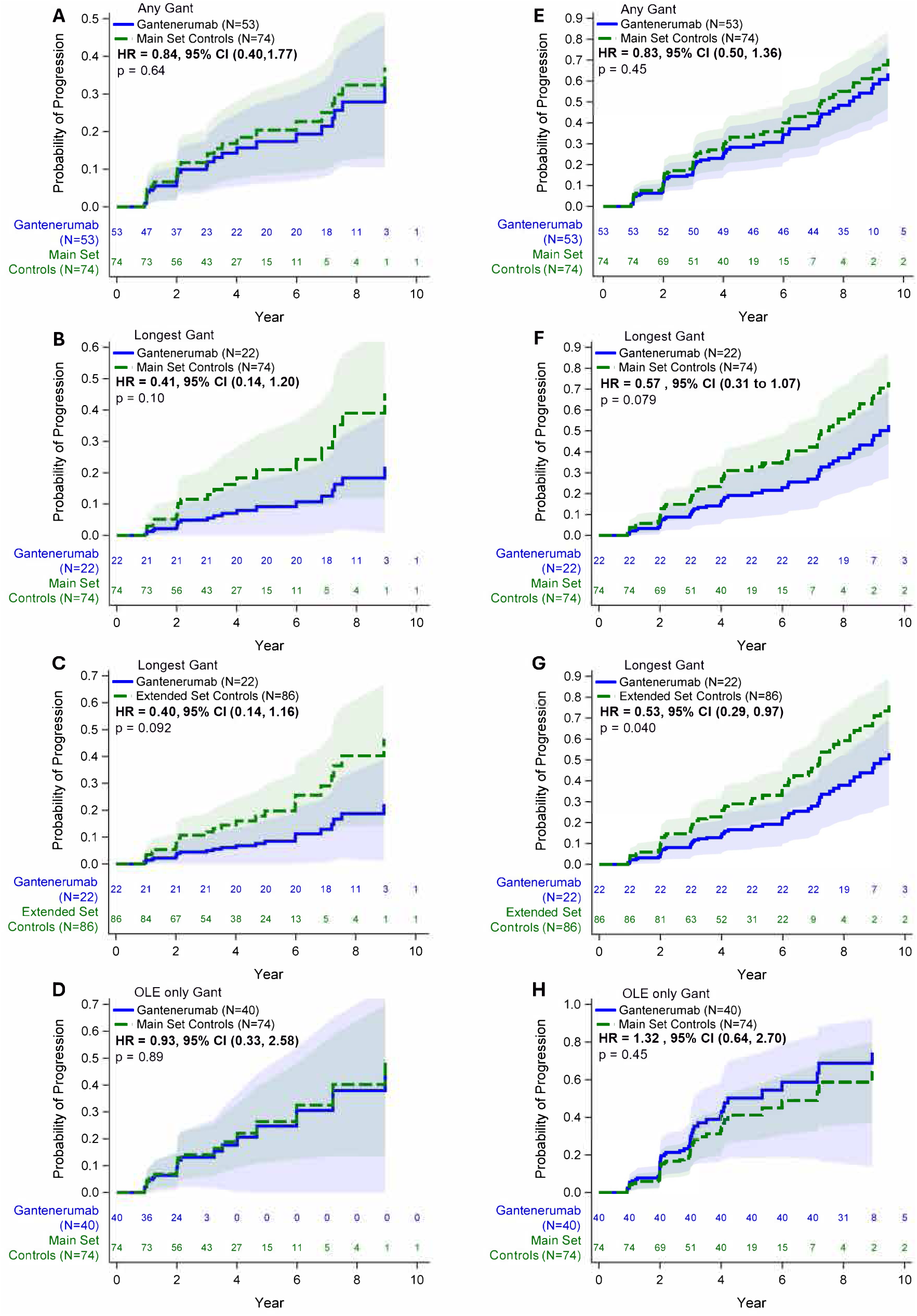
Probability of progression in CDR global and CDR-SB estimated from the Cox proportional hazards model. Lower probability of progression in treatment group and hazard ratio indicates higher reduction in the risk of progression and therefore better treatment effect. A and B. Any Gant mITT population baseline asymptomatic group vs main set of control; C and D. Longest Gant mITT population baseline asymptomatic group vs main set of control; E and F. Longest Gant mITT population baseline asymptomatic group vs extended set of control; G and H. OLE only Gant mITT population baseline asymptomatic group vs main set of control.

For the subset of baseline asymptomatic participants with the Longest Gant treatment (n=22, average 8.44 years), the hazard ratio (95% CI) of time to first progression in CDR global for treatment group (n=22) compared to the main set of controls (n=74) was 0.41 (0.14 to 1.20); that for time to recurrent progression in CDR-SB was 0.57 (0.31 to 1.07). When comparing the treatment group with the extended control group (n = 86), the hazard ratio (95% CI) is 0.40 (0.14 to 1.16) for time to first progression in CDR global and 0.53 (0.29 to 0.97) for time to recurrent progression in CDR-SB (Figure 3, Supplemental Table 3). For each population, the number and percentage of censored participants and reasons for censoring are presented in Supplemental Table 4. Sensitivity analyses conducted either by excluding double-blind placebo arm participants who did not enroll in OLE from the main set of control or by using control participants matched by baseline EYO, length of follow-up and mutation type, resulted in similar hazard ratios (Supplemental Table 5).

For the OLE baseline asymptomatic group and the OLE period data only, the hazard ratio (95% CI) of time to first progression in CDR global for treatment group (n = 40) compared to the main set of control (n=74) was 0.93 (0.33 to 2.58) and 1.32 (0.64, 2.70), respectively, for time to recurrent progression in CDR-SB.

### Other secondary outcomes: cognitive and biomarker

The Functional Assessment Scale, MMSE, and OLE cognitive composite outcomes showed no significant differences in the annual rate of change between the gantenerumab treated group (OLE period data only) and the main set of controls for either the OLE baseline asymptomatic group or baseline symptomatic group (Table 2). Post-hoc analyses of recurrent progression in FAS and MMSE for baseline asymptomatic participants with the Longest Gant treatment compared to the extended set control showed the hazard ratio was 0.59 (95% CI 0.22 to 1.58, p=0.29) for FAS and 0.54 (95% CI 0.21 to 1.39, p=0.20) for MMSE.

**Table 2.**
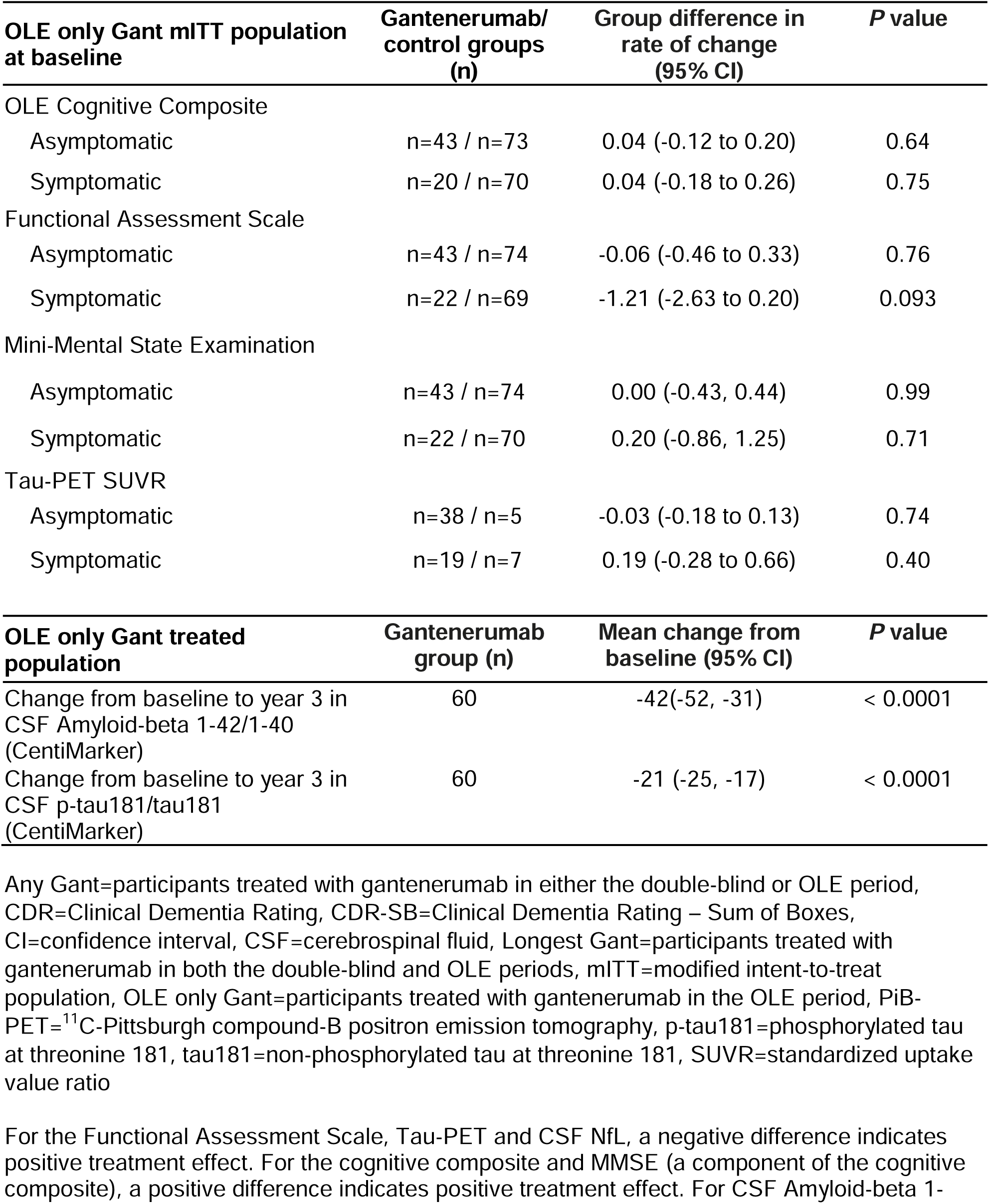
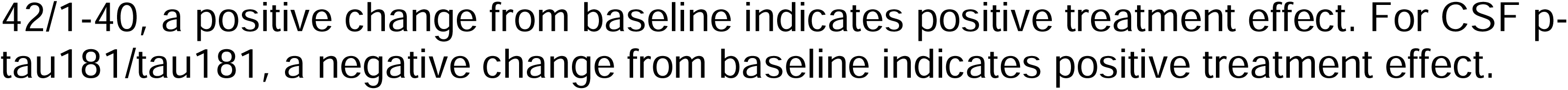
Final Analysis Secondary outcomes. For cognitive composite, FAS and MMSE, positive difference in rate of change indicates positive treatment effect. For tau PET SUVR, negative difference in rate of change indicates positive treatment effect. For CentiMarker, negative change from baseline indicates positive treatment effect.

Gantenerumab treatment during the OLE period significantly improved CSF Aβ 1-42/1-40 (mean change from OLE baseline at year 3 in CentiMarker: -42; 95% CI, -52 to -31, p<0.0001) and improved CSF p-tau181/tau181 (mean change from OLE baseline at year 3 in CentiMarker: -21; 95% CI, -25 to -17, p<0.0001) (Table 2 and Figure 2B,C). Change from baseline in original scale were provided in Supplemental Table 9.

No significant differences in the annual rate of change of Tau-PET SUVR were observed between the treatment group (OLE period only) and the internal control group for either the OLE baseline asymptomatic group or the symptomatic group (Table 2).

### Safety

The safety profile of gantenerumab at doses up to 1500 mg every 2 weeks was consistent with that observed during the double-blind period^17^ and in other trials in SAD populations.^12^ However, the ARIA-E rate was approximately one-third higher in the OLE compared to the rate observed with the lower doses in the double-blind period. Table 3 and Supplemental Table 6 summarizes the incidence of TEAEs, which were greater than 10% in the OLE period. The most common TEAEs were infusion-related reactions (70%), upper respiratory tract infections (59%), and upper respiratory symptoms (45%). Treatment-emergent serious adverse events (SAEs) and adverse events leading to treatment discontinuation are detailed in Table 3 and Supplemental Table 6, summarized by system organ class and preferred terms. Among the 73 participants, 71 participants had at least on TEAE, 9 participants (12%) experienced SAEs with 4/9 within the category of nervous system disorders. Two participants (3%) discontinued treatment and study due to AE. There was no death during the OLE period.

**Table 3.** Treatment Emergent Adverse Events by Treatment Group.

| Double-blind period |  |  | Open-label extension period |  |  |
| --- | --- | --- | --- | --- | --- |
|  | Shared placebo<br>(N=40)<br>n (%) | Gantenerumab<br>(N=52)<br>n (%) |  | Gantenerumab<br>(≥1020 mg Q2W; N=60)<br>n (%) | Gantenerumab<br>(all doses; N=73)<br>n (%) |
| Injection site reactions | 18 (45) | 47 (90) | Infusion<br>reactions | 47 (78) | 51 (70) |
| Amyloid related imaging abnormalities (ARIA) |  |  |  |  |  |
| ARIA-E | 1 (3) | 10 (19) |  | 17 (28) | 22 (30) |
| ARIA-H |  |  |  |  |  |
| Microhemorrhage | 4 (10) | 13 (25) |  | 24 (40) | 34 (47) |
| Superficial<br>siderosis | 0 (0) | 2 (4) |  | 3 (5) | 5 (7) |

ARIA=amyloid-related imaging abnormalities, ARIA-E=amyloid-related imaging abnormalities with edema, ARIA-H=amyloid-related imaging abnormalities with haemosiderosis/microhemorrhages, LP=lumbar puncture, Q2W=every 2 weeks

During the OLE period, the combined incidence of amyloid-related imaging abnormalities (ARIA), including both ARIA-E (cerebral edema/effusion) and ARIA-H (hemorrhage), was 53% (39/73). Specifically, the incidence of ARIA-E was 30% (n=22/73). Among those receiving higher doses (≥1020 mg every 2 weeks), the incidence of ARIA-E was 28% (n=17/60). Additionally, eight out of 22 participants experienced recurrent ARIA-E, totaling 32 episodes. Most ARIA-E episodes occurred within the first 4.7 months of receiving the higher doses. MRI analyses show that most ARIA-E events were multifocal, had a predominant occipital distribution, and a mean resolution time of 59.5 days. ARIA-E episodes were primarily asymptomatic (26/32). Symptomatic cases exhibited mild symptoms such as headache or transient disorientation. Symptomatic ARIA-E leading to hospitalization (worsening headaches and cortical blindness) occurred in two participants and both resolved with drug discontinuation. The frequency of ARIA-E was higher among *APOE* ε4 homozygous participants (2/3) compared to heterozygous participants (26%, 5/19) or non-carriers (29%,15/51). No cerebral macrohemorrhages were observed. No deaths were attributed to ARIA events.

## DISCUSSION

The amyloid hypothesis of AD pathophysiology predicts that removal of cerebral amyloid should prevent or delay the onset of dementia or slow cognitive-behavioral decline. Clinical trials in DIAD presents an opportunity to test the hypothesis by initiating therapy in persons with brain amyloid deposits prior to symptom onset. The DIAN-TU-001 double-blind and OLE trials provide the longest test of this approach, and results suggest that removal of brain amyloid plaque years before clinical symptom onset might be associated with delay in symptom onset.

There was no significant effect on the interim primary outcome of symptom onset CDR hazard ratio of shortest treated group (2-3 years, OLE only Gant) and the combined longest and shortest treated group (Any Gant). Further, no significant treatment effect was observed in the cognitive composite, MMSE and FAS. However, the outcomes from the longest-treated group (average 8.4 years, Longest Gant) suggest a possible benefit. The magnitude of effect of an approximately 50% slowing of dementia progression in the longest treated group is supported by multiple sensitivity analyses matching external and internal controls, the high predictability of disease onset and progression in DIAD mutation carriers, the large effect size point-estimate, similar magnitudes of effect on other clinical measures such as the FAS and MMSE, and the robustness of the CDR as a clinical measure of cognition and function. Further, ad-hoc analysis of the randomised controlled trial baseline asymptomatic gantenerumab arm double-blind period only, demonstrated a similar, but not significant, 49% slowing in the recurrent progression of CDR-SB compared to baseline asymptomatic placebo arm. These preliminary findings should be tested and confirmed with continued long-term follow-up and treatment of this unique cohort.

Even though the hazard ratios (HR) for CDR global, FAS and MMSE were not statistically significant, the magnitude of ∼40-50% are similar to HRs based on CDR-SB. For CDR global, only the first progression was considered and therefore it is less powerful than the recurrent progression for CDR SB. The negative findings of the Any Gant (the shortest and longest treated groups combined) and OLE cohorts in CDR, cognitive composite, MMSE and FAS based on OLE only period data could be due to double-blind solanezumab and placebo treated participants having only been treated for 2 to 3 years of gantenerumab without sufficient amyloid plaque removal to normal levels. Further, long-term follow-up may be needed, as only those followed for years can an effect in controls and treated groups be observed. Due to the limited sample size in this rare disease population, achieving statistically significant results is challenging and we note the magnitude of the effect sizes and 95% CI can provide biological and clinical information.

There was substantial dose-dependent amyloid removal as measured by the final primary outcome and analyses of dose-response. We tripled the dose of gantenerumab in the OLE study to increase amyloid removal. Even at this dose, amyloid plaques were not fully cleared in 70% of participants after one year of dose titration and one year at maximum doses, though there was substantial progress toward amyloid removal (most with Centiloid <25, supplemental figure 6) with an acceptable safety profile. Whether even more effective amyloid removal would further increase clinical benefit remains an open question.

In the presence of amyloid removal therapy, we observed improvement of biomarkers CSF Aβ 42/40 ratio and p-tau181, approaching or even reaching the values considered normal in control groups. In this trial, biomarker levels reached normal levels in asymptomatic participants, including in some individuals who were past their EYO, with a rate of biomarker change directly proportional to dosage, suggesting a potential dose-dependent effect, with a hypothetical plateau in the biomarker improvement towards normality when all plaques are removed. There was no significant treatment effect on tau PET in the overall group, similar to observations in SAD. However, future comparisons of the amount of tau pathology between the longest treated and control groups are underway.

The safety of anti-amyloid monoclonal therapy in early-stage symptomatic clinical AD (MCI and mild AD dementia) has been assessed in the context of restricted patient selection (e.g. microhemorrhages <5 with durations of only a few years). The participants in the DIAN-TU-001 trial have experienced the longest exposures to, and highest dosages of, gantenerumab administered to date, providing invaluable safety experience. Safety data from the DIAN-TU-001 OLE study indicate that tripling the dose of gantenerumab is reasonably well tolerated in DIAD populations, with the incidence and severity of adverse events remaining comparable to sporadic AD populations. However, higher doses in the OLE resulted in a third higher incidence than the double-blind period.^10^ Development of spontaneous ARIA with haemosiderosis or microhemorrhage (ARIA-H) and a potentially associated increasing risk of ARIA-E has been noted in this DIAD population, with ARIA most often seen in symptomatic participants or those nearing their mutation EYO. As trials are implemented in asymptomatic participants to treat earlier phases of amyloid plaque deposition or for the primary prevention of plaques, it will be important to learn the degree of risk for ARIA events associated with these earlier regimens. Our experience to date is consistent with reduced risk at earlier stages of amyloid pathology. Most ARIA events were not associated with clinical symptoms. Careful MRI monitoring— particularly in the early stages of anti-amyloid treatment—allowed therapy to proceed without evidence of lasting impairment caused by ARIA. Additionally, the higher doses of gantenerumab were not associated with new or unexpected safety findings. These safety results were obtained by using a slow dose escalation that also reduced the reaching and duration of exposure to full doses of gantenerumab, but may have allowed early detection and management strategies to closely assess (radiologically and clinically) and to mitigate progression and clinical impact of ARIA-E. This approach could have mitigated the severity and impact of ARIA, contributing to the relatively low number of symptomatic cases and ensuring that episodes were fully reversible. Alternatively, the slow titration may not have had an impact on the rate of ARIA, or ARIA detection, management, and safety outcomes.

Risks for ARIA-E in DIAD parallel those reported for gantenerumab in SAD, including increased risk of ARIA-E with *APOE* ε4 status and number of microhemorrhages.^12^ Observed ARIA-E rates were comparable to the TRAILBLAZER-ALZ2 and GRADUATE trials (∼25%) with gantenerumab in SAD. However, rates were higher than the 12% reported in the CLARITY AD trial of lecanemab in SAD. The higher ARIA-H relative to the double-blind period likely results from the combined effect of higher doses and disease progression. Previous DIAN Obs natural history studies have shown increased number and frequency of microhemorrhages with disease progression.^18^ Thus, the natural increase in vascular amyloid due to disease progression, combined with higher amyloid-modifying therapies doses, contributes significantly to the increased ARIA-H incidence during the OLE phase.

These results are encouraging but limited by the long asymptomatic phase of the disease, the incomplete removal of amyloid with gantenerumab, limited participant numbers, drop-outs, and the evolving nature of the trial design (Figure 1). The nature of OLE may introduce potential measurement and selection bias. To minimize bias, participants and assessors were blinded to the prior treatment assignment. A sensitivity analysis for a matched control group to account for potential baseline differences and selection bias was performed. These sensitivity analyses support the main findings of reduced clinical onset and progression of the longest treated group. We also evaluated attrition and survivorship bias, noting very little attrition in the asymptomatic group from baseline double-blind to the OLE period (28% total attrition). As the hazard ratio may change over time, its interpretation should be restricted to the follow-up duration of the study. The use of external and internal controls, the limited follow-up duration, and difference in the follow-up duration between treatment and control groups limit the conclusiveness of these observations, warranting additional follow-up. These limitations are mitigated by the large degree of amyloid plaque removal, the consistency between clinical and biological measures improving towards normal, and the large clinical effect size seen in the longest treated group.

Anti-amyloid therapy trials in presymptomatic populations at risk for developing AD, including the Alzheimer’s Prevention Initiative Autosomal Dominant Alzheimer’s Disease Colombia trial of crenezumab in individuals with *PSEN1* E280A mutation and the Anti-Amyloid Treatment in Asymptomatic Alzheimer’s Disease (A4) Phase 3 trial of solanezumab, have not demonstrated slowing of cognitive decline before. Amyloid removal measured by amyloid PET was minimal in each trial. Negative trial outcomes may be due to incorrect targets (e.g. soluble only species), inadequate doses or insufficient treatment duration (4 years for double-blind period vs. 8 years follow-up in this OLE). In this extended trial, we found clinical indicators of delayed progression in those treated for the longest with gantenerumab, which removed most amyloid plaques. To date, all trials in mutation carriers have predicted outcomes in late-onset SAD (see Supplemental Table 7), and so an open question is whether these findings will be replicated in ongoing prevention trials of SAD that remove plaques.^19,20^

In conclusion, while further investigation and larger-scale trials are needed to confirm the observed clinically significant but statistically uncertain clinical effects observed in the those treated the longest, we believe that amyloid removal (the final primary outcome) and biomarker effects offer cautious optimism that disease modification in individuals at risk for AD may be possible. We launched the DIAN-TU Amyloid Removal Trial (NCT06384573) to continue following this cohort to determine the robustness of these findings and if this cohort continues to be protected from dementia onset. Furthermore, ongoing follow-up could enable the development of more accurate ARIA predictors, which is critical for improving participant management, by identifying high-risk participants and increasing the frequency of monitoring and dose adjustments. Continued follow-up of this unique DIAN-TU cohort exposed to long-term amyloid-plaque reducing monoclonal antibodies, other DIAN-TU cohorts, and the results of other ongoing prevention trials in SAD will be crucial in shaping the future direction of AD prevention efforts.

### Research in context

#### Evidence before the study

In 2012, the Dominantly Inherited Alzheimer Network Trials Unit (DIAN-TU) launched a pioneering prevention platform trial (DIAN-TU-001, NCT01760005) to investigate the efficacy of anti-amyloid antibodies solanezumab and gantenerumab in delaying symptom onset and slowing progression in asymptomatic and mildly symptomatic Dominantly Inherited Alzheimer’s Disease (DIAD) mutation carriers. Other prevention trials launched and completed, including the Alzheimer’s Prevention Initiative (API) Autosomal Dominant Alzheimer’s Disease (ADAD) Colombia trial of crenezumab in 2013 (NCT01998841) and the Anti-Amyloid Treatment in Asymptomatic Alzheimer’s Disease (A4) trial of solanezumab in 2014 (NCT02008357). In the DIAN-TU-001 trial, gantenerumab did not meet its primary endpoint of slowing cognitive decline in combined cohorts that included participants with symptoms of dementia. However, it demonstrated significant but incomplete reduction of amyloid-plaques in a dose-dependent fashion, with improvements in CSF biomarkers of amyloid deposition, phosphorylated-tau, and neurodegeneration, particularly in asymptomatic DIAD mutation carriers. Furthermore, downstream CSF and blood-based biomarkers showed that gantenerumab treatment significantly improved multiple measures of synaptic degeneration, microglial activity, other inflammatory markers and astrocyte activation markers. In contrast, the prevention trials of solanezumab and crenezumab, antibodies targeting soluble amyloid-beta, did not demonstrate delay or slowing of cognitive decline or substantial amyloid removal measured by amyloid PET or improvements in other biomarkers. Together, these results provided the rationale for continuing gantenerumab treatment in an open-label extension study (NCT06424236) at higher doses to achieve full amyloid removal and determine if the onset of symptomatic disease progression could be delayed.

The evidence gathered to provide rationale for the study was from known literature of Alzheimer’s disease searched for on PubMed, prevention trials registered at ClinicalTrials.gov, and other repositories. The DIAN Observational Study, established in 2008, serves as the key scientific and discovery study for DIAD and provides a wealth of natural history information about clinical, cognitive, and biomarker trajectories in both the asymptomatic and symptomatic disease phases.^9,26^ These data provide the basis for DIAN-TU prevention trial design.^27,28^

### Added value of the study

The DIAN-TU gantenerumab findings suggest for the first time in a clinical trial, that early treatment and removal of amyloid plaques during the asymptomatic phase of disease might delay the onset and decrease the risk of AD progression. Furthermore, this study provides valuable insights into the safety of long-term treatment (∼8 years) with amyloid removing therapies at high doses.

### Implications of all the available evidence

Phase 3 trials in sporadic late-onset Alzheimer’s disease demonstrate treating early symptomatic stages (MCI to mild dementia) slows progression by ∼30%. However, the optimal timing to treat asymptomatic or presymptomatic individuals to prevent or delay Alzheimer’s remains uncertain. The DIAN-TU trial of gantenerumab suggests that substantial amyloid plaque removal in presymptomatic DIAD mutation carriers might delay and slow the onset and progression in the longest treated group by 50%, but only in those reaching near normal amyloid levels or treated many years before symptom onset. Further research is needed to clarify the extent to which complete amyloid removal can delay, slow, or prevent AD dementia in this group of DIAD mutation carriers and in the general population. These questions will be addressed in ongoing studies in DIAD (the DIAN-TU Amyloid Removal Trial), and in ongoing late-onset prevention trials.

## Data Availability

See pdf of manuscript.

## Contributors

R.J.B., Y.L., E.M.M., J.J.L.G., D.B.C., S.L.M., A.M.S., G.W., C.S., T.L.S.B., B.A.G., L.I., G.K., M.B., R.S.D., A.A., P.D., G.A.K., J.H., A.J.A., R.J.P, C.C., A.R., C.X. designed the study and wrote the study protocol. T.B., J.W., A.B., P.F., C.H., L.K., A.A.G., J.C.M., D.M.H. provided additional input to the study design. R.J.B., E.M.M., D.B.C., J.J.L.G, S.L.M. and A.M.S. coordinated the trial. B.J.S., C.M., D.W., S.B.B., E.D.R., C.H.V.D., R.S., W.S.B., S.G., D.R.G., C.L.M., S.J., G.R.H., M.M., B.D, J.P., J.K., L.H. recruited and treated the patients and collected the data. G.W. and Y.L. accessed and verified the data. G.W., Y.L., C.X., T.B., and P.D. performed the statistical analysis. S.L.M. and A.M.S. were responsible for central and local data management. L.I. performed the analysis of the CSF biomarkers data. C.R.J., T.L.S.B. and B.A.G. performed the processing and analysis of the neuroimaging data. C.C. and A.R. were responsible for the sequencing and genetic analysis. Every author contributed to the initial draft of the manuscript and agreed on submission for publication. All authors interpreted the data, reviewed the manuscript and approved the final version. All authors had full access to all the data in the study and were responsible for the decision to submit for publication.

## Data Sharing

Data access to the DIAN–TU trial data will follow the policies of the DIAN–TU data access policy (https://dian.wustl.edu/wp-content/uploads/2021/02/DIANTUResourceSharePublAuthorPolicy_Ver1.0_12Jan2021.pdf), which complies with the guidelines established by the Collaboration for Alzheimer’s Prevention. Patient-related data not included in the paper were generated as part of a clinical trial and may be subject to patient confidentiality. Any data and materials that can be shared will be released via a data/material sharing agreement. Requests to access the DIAN–TU-001 trial data can be made at https://dian.wustl.edu/our-research/for-investigators/diantu-investigator-resources/. All code for data cleaning and analysis associated with the current submission is available upon request to the corresponding author and is provided as part of the replication package.

## Acknowledgements

Research reported in this publication was supported by the National Institute on Aging of the National Institutes of Health under Award Numbers U01AG042791, U01AG042791-S1 (FNIH and Accelerating Medicines Partnership), R1AG046179, R01AG053267, R01AG053267-S1, R01AG053267-S2. The research for the DIAN-TU-001 gantenerumab open label extension was supported by the Alzheimer’s Association and F. Hoffman-LaRoche Ltd. The research for the DIAN-TU-001 trial, solanezumab and gantenerumab drug arms, was also supported by the Alzheimer’s Association, Eli Lilly and Company, F. Hoffman-LaRoche Ltd., Avid Radiopharmaceuticals (a wholly owned subsidiary of Eli Lilly and Company), GHR Foundation, an anonymous organization, Cerveau Technologies, Cogstate, and Signant. The DIAN-TU has received funding from the DIAN-TU Pharma Consortium. We acknowledge the altruism of the participants and their families and contributions of the DIAN, DIAN Expanded Registry, and DIAN-TU research and support staff at each of the participating sites (see DIAN-TU Study Team) for their contributions to this study.

The content is solely the responsibility of the authors and does not necessarily represent the official views of the National Institutes of Health.

Statistical data analysis support was provided by Daniel Yen.

