## Supplemental Materials for "Safety and efficacy of long-term gantenerumab treatment in dominantly inherited Alzheimer’s disease: an open label extension of the phase 2/3 multicentre, randomised, double-blind, placebo-controlled platform DIAN-TU Trial"

### SUPPLEMENTARY FIGURES, TABLES AND METHODS

Supplemental Table 8. Conversion from abnormal to normal amyloid-plaque status from gantenerumab OLE baseline to the last post-baseline OLE visit in PiB-PET status based on OLE only Gant mITT population. . ..........................................................................16

**Supplemental Method 1. Post-hoc sensitivity analysis for time to first progression in CDR global and the time to recurrent progression in CDR-SB.**

As the Longest Gant mITT population only included participants who enrolled in OLE while the main set of control included double blind placebo group participants who did not enroll in OLE, a sensitivity analysis was performed by excluding double blind placebo participants who didn't enroll in OLE from the main set of controls. Then the same Cox proportional hazards model as described in the statistical method section was used to estimate the hazard ratio.

Another sensitivity analysis was performed by selecting a matched subset of participants from the extended set of control using propensity score matching. The matching was based on the nearest logistic regression propensity scores of baseline EYO and mutation status (*PSEN1*, *PSEN2* and *APP*). Propensity score matching was performed using R function matchit with caliper of 0.2 (standard deviation of the propensity score). The same Cox proportional hazards model as described in the statistical method section but with baseline EYO and mutation status as strata was then applied to the matched dataset to estimate the hazard ratio.

**Supplemental Methods 2. Methods**

**Procedures**

*Assessments*

Clinical and cognitive assessments were performed at baseline and every 6 months. These included CDR global and Sum-of-Box scores (CDR-SB), Functional Assessment Scale (FAS), Mini-Mental State Examination (MMSE), and the DIAN-TU OLE Cognitive Composite (consisting of 4 components: the Wechsler Memory Scale-Revised Digit Span Backward Recall, the Category Fluency (Animals) total correct score, the Wechsler Adult Intelligence Scale Digit Symbol Substitution Test total correct score, and the total score from the MMSE, Supplemental Methods 2). Blood collection for serum and/or plasma was performed every 6 months. Volumetric MRI (vMRI), $^{11}$C-Pittsburgh compound-B positron emission tomography (PiB-PET), flortaucipir (Tauvid®) PET (Tau-PET) and lumbar puncture for CSF collection were conducted annually. In addition to the annual vMRIs, safety surveillance MRIs for amyloid-related imaging abnormalities (ARIA)[21] monitoring were collected at 9 weeks, then every 12 weeks thereafter. Safety follow-up visits were performed 12 weeks after the last dose of gantenerumab. Safety assessments included adverse events, pregnancy testing and vitals at each dosing visit, and routine laboratory assessments and physical examinations every 6 months.

*Imaging methods*

Structural MRI was performed using the Alzheimer's Disease Neuroimaging Initiative (ADNI) protocol using 3T MRI scanners. The ADNI Imaging Core screened images for protocol compliance, artifacts and ARIA. T1-weighted images (1.1 × 1.1 × 1.2 mm voxels) were acquired for all participants as to perform volumetric quantification. The presence and number of cerebral microbleeds and siderosis was evaluated on gradient echo images (GRE, 0.86 x 0.86 x 4 mm voxels). ARIA-E was evaluated on FLAIR images (0.86 x 0.86 x 5.0 mm voxels).
Regions of interest (ROIs) were defined by FreeSurfer on the MRI scans were used for the regional processing of all PET data. PiB-PET data from the 40–70-min postinjection window were converted to regional standard uptake value ratios (SUVRs) relative to the cerebellar gray

matter (https://github.com/ysu001/PUP). A composite to represent a global measure of Aβ-plaques was calculated using the averaged SUVR values in the precuneus, caudate, gyrus rectus, occipital cortex, parietal cortex, prefrontal cortex and temporal cortex regions. The Tau-PET summary region is the average SUVR values for the postcentral gyrus, supramarginal gyrus, superior parietal cortex, inferior parietal cortex, precuneus, posterior cingulate, and isthmus cingulate. Both types of PET data were partial volume-corrected using a regional spread function technique.[22]

*Fluid biomarker methods*

Cerebrospinal fluid was collected at DIAN-TU sites under fasting conditions using a 22-G atraumatic Sprotte spinal needle (except when fluoroscopy was required) into a single 50 mL polypropylene tube via gravity drip methods. The fluid was immediately processed into 10 0.5 mL aliquots in 2.0 mL polypropylene tubes and flash frozen. The tubes were shipped overnight on dry ice to the central laboratory (LabCorp) for storage at −80 °C. Samples were shipped to the DIAN-TU Biomarker Core laboratory at Washington University, St. Louis, Missouri for analysis. All tubes were stored at −80 °C.

One freeze-thaw cycle CSF aliquots were used for the analysis of Aβ1-40, Aβ1-42, and p-tau concentrations measured using high-precision immunoprecipitation-coupled mass spectrometry (IP-MS) as previously described.[23,24]

**Statistical Analysis**

*Power Calculations*

The OLE study successfully enrolled 74 participants and dosed 73 (1 participant screen failed). We estimated annual dropout rates of 5% and 10% and anticipated approximately 56 to 43 participants to remain at the end of the 3-year OLE. The power estimation was based on the primary efficacy endpoint of the change from OLE baseline to the end of the OLE period. Based on the gantenerumab treatment effect observed during double blind period,[10] the mean (SD) change from OLE baseline in PiB-PET SUVR was projected to be at least -0.47 (0.54) over the 3-year OLE period, with dosing up to 1500 mg every 2 weeks (Supplemental Method 5). Given the anticipated sample size, the study was predicted to have over 99% power to detect the projected mean change from baseline using a one-sided one-sample t-test with a type I error rate of 3%. One-sided test was used since double-blind period data already demonstrated gantenerumab can significantly reduce PiB-PET SUVR from baseline.

**Supplemental Method 3. DIAN-TU Cognitive Composite Calculation**

The DIAN-TU Cognitive Composite which includes the Wechsler Memory Scale-Revised (WMS-R) Digit Span Backward Recall, the Category Fluency (Animals) value, the Wechsler Adult Intelligence Scale Digit Symbol Substitution Test value, and the Mini-Mental State Examination (MMSE) calculated using the following formula:

$$Y=(0.25)\frac{M-mean}{SD}+(0.25)\frac{W-mean}{SD}+(0.25)\frac{A-mean}{SD}+(0.25)\frac{D-mean}{SD}$$

where, D represents the WMS-R Digit Span Backward Recall, A represents the Category Fluency (Animals) value, W represents the Wechsler Adult Intelligence Scale Digit Symbol

Substitution Test value, and M represents the MMSE value and the mean (SD) for each of the four parameters were calculated using the double-blind baseline data of the mutation carriers.

**Supplemental Method 4. Interim Analysis**

The two objectives of the interim analysis were to determine if gantenerumab OLE treatment resulted in clinical benefit and determine the extent of amyloid removal compared to the double-blind period in participants who were asymptomatic at baseline compared to main set controls (external controls and placebo data from participants in the double-blind period that did not enter the OLE). The pre-specified primary outcome measures for the interim analysis were the time to first progression of Clinical Dementia Rating (CDR) Global and recurrent progression of CDR – Sum of Boxes (CDR-SB). Key secondary biomarker measure for amyloid deposition is a composite PiB partial volume corrected standardized uptake value ratio (PiB-PET SUVR) which is a composite SUVR of the precuneus, caudate, gyrus rectus, occipital cortex, parietal cortex, prefrontal cortex and temporal cortex regions, with values expected to decrease with effective drug.

**Supplemental Method 5: Preliminary data used for power analysis.**

Based on double-blind period data, the mean annual reduction in PiB-PET SUVR was estimated to be 0.05 for low dose period and 0.105 for high dose period (1200 mg every 4 weeks).[10] The projection for mean change from OLE baseline to OLE year 3 assumes the SUVR reduction during the first year of OLE is similar to the low dose period in double-blind phase and the SUVR reduction during OLE year 2 and 3 (1020~1500 every 2 weeks) is twice that in double-blind high dose period since the dose in OLE is more than doubled. The SD estimated based on the double-blind period year 4 variance from covariance matrix of MMRM is 0.54. This is a conservative estimation since the OLE period is only 3 years and SD could be smaller.

**Supplemental Method 6. CentiMarker Calculation**

In order to convert the raw biomarker values to CentiMarker values, we utilized a similar methodology as the Centiloid conversion approach. For CentiMarker calculations, more abnormal biomarker raw values indicate worse disease stages as defined by the direction of disease vs. normal groups. In summary, the process involves the following steps:

1. To establish the CentiMarker-0 anchor ($\mu_{\mathrm{CM-0}}$), all the data from the asymptomatic mutation non-carriers enrolled in the DIAN-TU trial is utilized. The first step involves calculating the interquartile range (IQR) of the data. Then, any outliers that fall outside the range of ($> \mathrm{Q3} + 1.5 \times \mathrm{IQR}$) or ($< \mathrm{Q1} - 1.5 \times \mathrm{IQR}$) are excluded from the dataset. Finally, the mean of the remaining data points was established as the CentiMarker-0 anchor ($\mu_{\mathrm{CM-0}}$) for subsequent CentiMarker calculations.

2. To determine the CentiMarker-100 anchor ($\mu_{\mathrm{CM-100}}$), data from all mutation carriers enrolled in the DIAN-TU trial who did not receive active treatments were used. This includes both the baseline data from participants who were assigned to the active treatment (gantenerumab and solanezumab) as well as the baseline and post-baseline data from those who were assigned to placebo. The same approach applied in the identification of the CentiMarker-0 data set, is again employed to identify and remove outliers from this dataset. The CentiMarker-100 anchor, denoted as $\gamma_{\mathrm{MC-100}}$, is established as the 95th percentile of the most abnormal value across the spectrum of

disease stages in the CentiMarker-100 dataset. With this approach, when the higher values indicate more severe disease stage, the 95th percentile of the most abnormal value corresponds to the 95th percentile of the highest values; when the lower values indicate more severe disease stage, the 95th percentile of the most abnormal value corresponds to the 5th percentile of the lowest values.

3. The CentiMarker is then calculated as:

$$\mathrm{CM} = \frac{\mathrm{y} - \mu_{\mathrm{CM}-0}}{\gamma_{\mathrm{CM}-100} - \mu_{\mathrm{CM}-0}} \times 100$$

where $\mathrm{y}$ is the raw biomarker value, $\mu_{\mathrm{CM}-0}$ is the CentiMarker-0 anchor, $\gamma_{\mathrm{CM}-100}$ is the CentiMarker-100 anchor.

**Supplemental Figure 1. DIAN-TU-001 gantenerumab OLE dose titration and MRI monitoring schedule.**

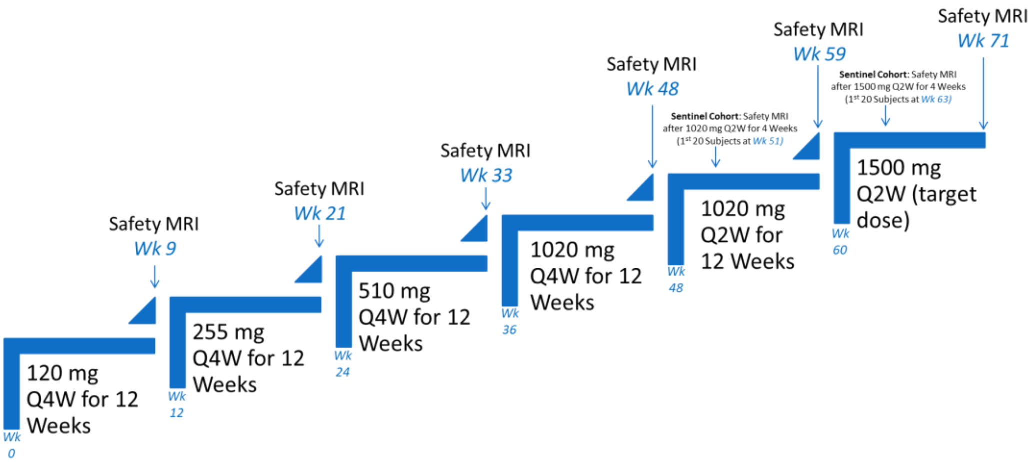

The titration schedule took place over 19 months with the target dose goal of 1500 mg gantenerumab administered every 2 weeks (Q2W), for a total of 3000 mg Q4W.

**Supplemental Table 1. Length of follow-up.**

| mITT Population | Baseline Group | Length of follow-up (years) Median (Q1, Q3) |
|---|---|---|
| Any Gant | All participants (n=93) | 3.19 (2.07, 6.98) |
| | Asymptomatic group (n=54) | 3.49 (2.71, 8.05) |
| | Symptomatic group (n=39) | 2.84 (1.99, 4.02) |
| Longest Gant | All participants (n=24) | 8.31 (7.83, 8.89) |
| | Asymptomatic group (n=22) | 8.25 (7.82, 8.98) |
| | Symptomatic group (n=2) | 8.48 (8.30, 8.65)* |
| OLE only Gant | All participants (n=65) | 2.64 (1.99, 2.90) |
| | Asymptomatic group (n=43) | 2.77 (1.99, 2.97) |
| | Symptomatic group (n=22) | 2.01 (1.98, 2.80) |
| Main set of control | All participants (n=144) | 3.21 (2.01, 4.35) |
| | Asymptomatic group (n=74) | 3.81 (2.19, 4.98) |
| | Symptomatic group (n=70) | 2.99 (1.26, 4.24) |
| Extended set of control | All participants (n=162) | 3.96 (2.05, 5.00) |
| | Asymptomatic group (n=86) | 3.99 (2.40, 5.65) |
| | Symptomatic group (n=76) | 3.05 (1.63, 4.69) |
| Main set of control without participants who did not enroll in OLE for sensitivity analysis | Asymptomatic group (n=76) | 4.13 (2.38, 5.89) |
| Matched control group by baseline EYO, length of follow-up and mutation type for sensitivity analysis. | Asymptomatic group (n=44) | 4.23 (3.97, 6.01) |

*Minimum and maximum are provided instead of Q1 and Q3 since there are only two participants.

**Supplemental Table 2. Primary analysis results from interim analysis.** Hazard ratio and *p* value are for comparisons between treatment group and control group. Lower hazard ratio indicates higher reduction in the risk of progression and therefore better treatment effect. Main set of control was used as the control group unless otherwise specified in the table.

| **Primary efficacy endpoint – baseline asymptomatic group mITT Population** | **Number of participants (treatment group/control group)** | **Hazard ratio (95% CI)** | ***P* value** |
|---|---|---|---|
| Time to first progression in CDR global | | | |
| Any Gant | n=53 / n=74 | 0.91 (0.43, 1.94) | 0.81 |
| Longest Gant | n=22 / n=74 | 0.43 (0.15, 1.26) | 0.12 |
| Longest Gant (vs extended control) | n=53 / n=86* | 0.42 (0.15, 1.22) | 0.11 |
| OLE only Gant | n=40 / n=74 | 1.39 (0.46, 4.19) | 0.56 |
| Time to recurrent progression in CDR SB | | | |
| Any Gant | n=53 / n=74 | 0.79 (0.47, 1.32) | 0.36 |
| Longest Gant | n=22 / n=74 | 0.53 (0.27, 1.03) | 0.060 |
| Longest Gant (vs extended control) | n=22 / n=86* | 0.49 (0.26, 0.94) | 0.032 |
| OLE only Gant | n=40 / n=74 | 1.41 (0.59,3.33) | 0.44 |

*Sample size for extended control.

**Supplemental Table 3. Primary and key secondary outcomes.** Lower hazard ratio indicates higher reduction in the risk of progression and therefore better treatment effect.

| **Primary efficacy endpoint** | **n** | **Mean change from baseline (95% CI)** | ***P* value** |
|---|---|---|---|
| Change from baseline to year 3 in PiB-PET composite – OLE only Gant mITT | 55 | -0.71 (-0.88 to -0.53) | <0.0001 |
| Change from baseline to year 3 in Centiloid – OLE only Gant mITT | 55 | -29 (-39 to -20) | <0.0001 |

| **Secondary efficacy endpoint - baseline asymptomatic group** | **Treatment group/ control group (n)** | **Event Rate per Person Year** | **Hazard ratio (95% CI)** | ***P* value** |
|---|---|---|---|---|
| *Time to first progression in CDR global* | | | | |
| Any Gant | n=53 / n=74 | 5.4%/5.9% | 0.84 (0.40 to 1.77) | 0.64 |
| Longest Gant | n=22 / n=74 | 3.0%/5.9% | 0.41 (0.14 to 1.20) | 0.10 |
| Longest Gant (vs extended control) | n=22 / n=86* | 3.0%/5.7% | 0.40 (0.14 to 1.16) | 0.092 |
| OLE only Gant | n=40 / n=74 | 7.6%/5.9% | 0.93 (0.33 to 2.58) | 0.89 |
| *Time to recurrent progression in CDR-SB* | | | | |
| Any Gant | n=53 / n=74 | 12.7%/10.6% | 0.83 (0.50 to 1.36) | 0.45 |
| Longest Gant | n=22 / n=74 | 9.7%/10.6% | 0.57 (0.31 to 1.07) | 0.079 |
| Longest Gant (vs extended control) | n=22 / n=86* | 9.7%/10.8% | 0.53 (0.29 to 0.97) | 0.040 |
| OLE only Gant | n=40 / n=74 | 15.8%/10.6% | 1.32 (0.64 to 2.70) | 0.45 |

*Sample size for extended control.

Any Gant=participants treated with gantenerumab in either the double-blind or OLE period, CDR=Clinical Dementia Rating, CDR-SB=Clinical Dementia Rating – Sum of Boxes, Longest Gant=participants treated with gantenerumab in both the double-blind and OLE periods, mITT=modified intent-to-treat population, OLE only Gant=participants treated with gantenerumab in the OLE period, PiB-PET=$^{11}$C-Pittsburgh compound-B positron emission tomography, SUVR=standardized uptake value ratio

**Supplemental Figure 2. Annual PiB-PET SUVR reduction by gantenerumab dose.**

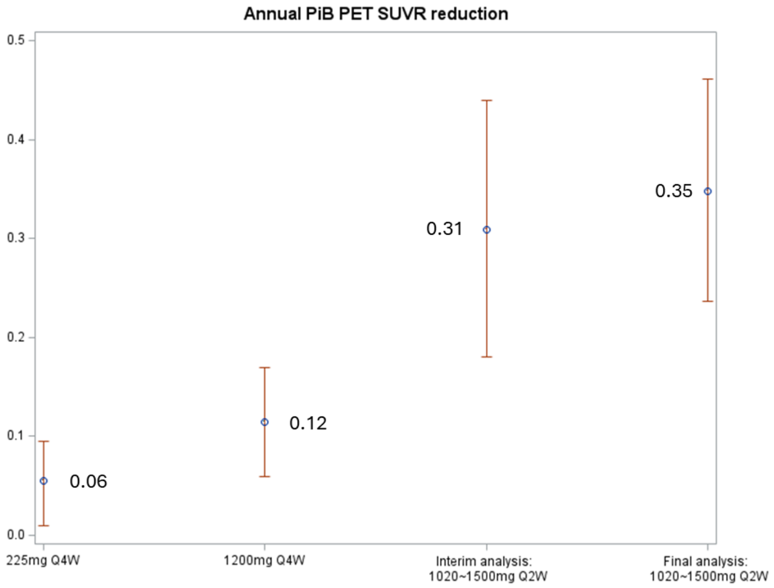

**Supplemental Figure 3. Gantenerumab treatment effects on PiB PET composite SUVR.** A. change from the gantenerumab open-label extension (OLE) baseline during OLE period; B. change from double-blind baseline during the whole study duration (DB + OLE) based on MMRM model. Solanezumab double-blind period data is not shown.

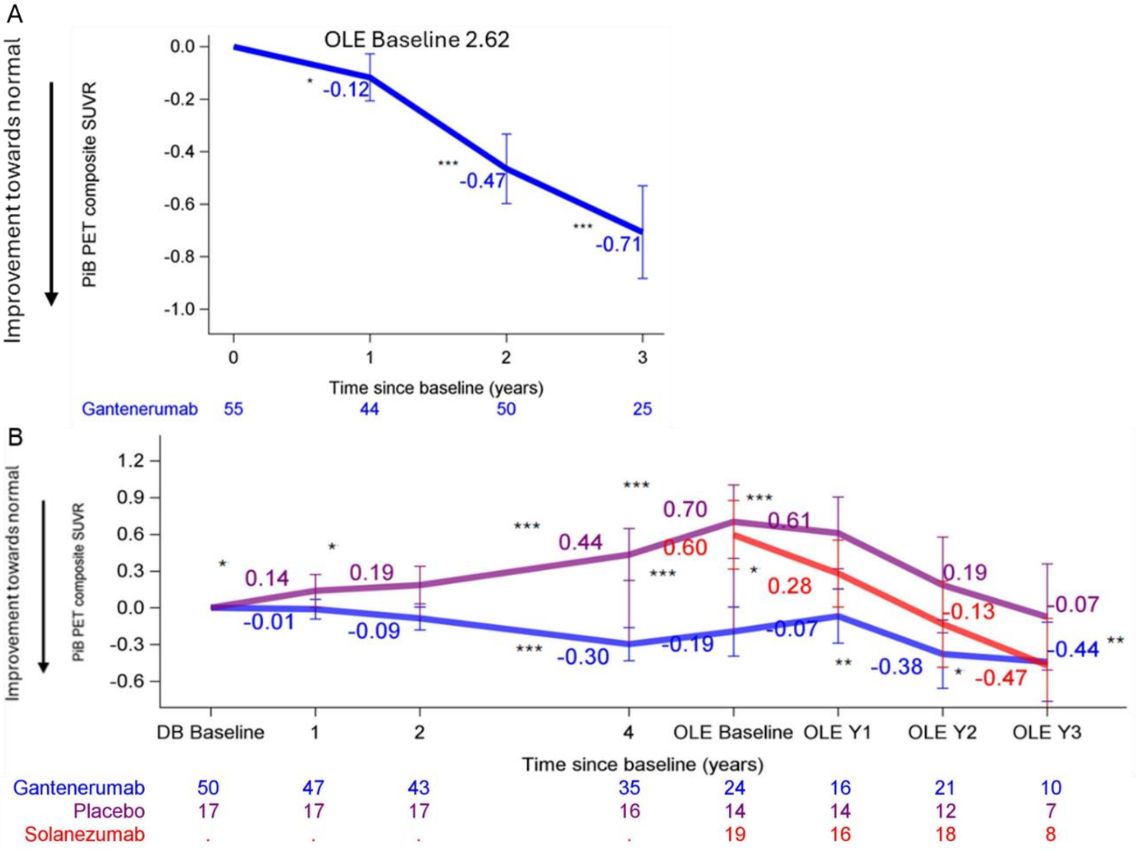

*P < 0.05, **P < 0.01, ***P < 0.001.

**Supplemental Table 4. Censored participants in the (A) treated groups and (B) controls.** Number (%) censored and reason for censoring in the analysis for time to current progression in CDR SB and time to first progression in CDR global.

**A.**

| **Reason for censor** * | **Any gantenerumab treated group (n = 53)**<br><br>**Censored: 37 (70%)** | **Longest gantenerumab treated group (n = 22)**<br><br>**Censored: 15 (68%)** | **OLE only gantenerumab treated group (n = 40)**<br><br>**Censored: 32 (80%)** |
|---|---|---|---|
| Completed study | 10 (27%) | 6 (40%) | 8 (25%) |
| Study terminated by sponsor | 24 (65%) | 9 (60%) | 24 (75%) |
| Non-compliance with study drug | 1 (3%) | 0 | 1 (3%) |
| Withdraw by subject or proxy | 2 (5%) | 0 | 2 (5%) |

**B.**

| **Reason for censor** | **Main control group (n = 74)**<br>**Censored: 56 (76%)** | **Extended control group (n = 86)**<br>**Censored: 65 (76%)** |
|---|---|---|
| Completed study | 41 (73%) | 50 (77%) |
| Elects to Enroll in a Clinical Trial | 6 (11%) | 6 (9%) |
| Lost to follow-up | 3 (5%) | 3 (5%) |
| Participant Burden | 2 (4%) | 2 (3%) |
| Consent Withdrawn | 2 (4%) | 2 (3%) |
| Adverse Event | 1 (2%) | 1 (2%) |
| Non-Compliance | 1 (2%) | 1 (2%) |

*Completed study included participants who either completed double-blind period of the study, but didn't enroll in OLE and participants who completed OLE.

**Supplemental Table 5. Sensitivity analyses results for CDR global and CDR-SB.** Data indicates that the magnitude of treatment effect did not substantially change when accounting for the differences in attrition, baseline EYO and mutation type.

| **Sensitivity analysis by excluding double-blind placebo arm participants who did not enroll in OLE from the main set of control** | | | |
|---|---|---|---|
| | **Hazard ratio** | **95% CI** | ***p*-value** |
| CDR Global | 0.52 | 0.18 to 1.54 | 0.24 |
| CDR SB | 0.57 | 0.30 to 1.07 | 0.079 |
| **Sensitivity analysis using control participants matched by baseline EYO and mutation type** | | | |
| CDR Global | 0.36 | 0.07 to 2.01 | 0.25 |
| CDR SB | 0.20 | 0.05 to 0.74 | 0.016 |

**Supplemental Table 6. Other adverse events reported**

| Double-blind period | | | Open-label extension period | | |
|---|---|---|---|---|---|
| | **Shared placebo (N = 40) n (%)** | **Gantenerumab (N = 52) n (%)** | | **Gantenerumab (≥1020 mg Q2W; N = 60) n (%)** | **Gantenerumab (all doses; N = 73) n (%)** |
| Injection site reactions | 18 (45) | 47 (90) | Infusion reactions | 47 (78) | 51 (70) |
| Nasopharyngitis | 11 (28) | 20 (38) | Upper respiratory tract infection | 38 (63) | 43 (59) |
| Back pain | 11 (28) | 16 (31) | Upper respiratory symptoms | 29 (48) | 33 (45) |
| Contact dermatitis | 5 (13) | 10 (19) | Headache | 22 (37) | 25 (34) |
| Nasal congestion | 4 (10) | 9 (17) | ARIA-E | 14 (23) | 19 (26) |
| Bronchitis | 5 (13) | 8 (15) | Post LP back pain | 18 (30) | 19 (26) |
| Muscle spasms | <4 (<10) | 8 (15) | Diarrhea | 10 (17) | 12 (16) |
| Sinusitis | 4 (10) | 8 (15) | Post LP syndrome | 9 (15) | 12 (16) |
| Fall | <4 (<10) | 6 (12) | Nausea | 9 (15) | 11 (15) |
| Oropharyngeal pain | <4 (<10) | 6 (12) | Arthralgia | 7 (12) | 9 (12) |
| | | | Fatigue | 8 (13) | 9 (12) |
| | | | Depression | 7 (12) | 8 (11) |
| | | | Pain in extremity | 7 (12) | 8 (11) |
| Post-baseline ARIA | | | | | |
| ARIA-E | 1 (3) | 10 (19) | | 17 (28) | 22 (30) |
| Microhemorrhage | 4 (10) | 13 (25) | | 24 (40) | 33 (45) |
| Superficial siderosis | 0 (0) | 2 (4) | | 3 (5) | 5 (7) |

**Supplemental Table 7. Dominantly inherited Alzheimer's disease trial outcomes are predictive of sporadic AD trials.**

| Antibody | Results of Sporadic AD Trials | Concordant results from DIAD Trials |
|---|---|---|
| Soluble Aβ-antibody | **A4 secondary prevention:** Solanezumab had little effect on plaques, clinical trend towards worsening | **DIAN-TU secondary prevention:** Solanezumab: Little effect on plaques, clinical and biomarker outcome worsening |
| | **Expedition 3 symptomatic:** Solanezumab did not reduce plaque, did not show clinical benefit overall | **DIAN-TU symptomatic:** Solanezumab did not reduce plaque, no clinical and biomarker benefit |
| | **CREAD symptomatic:** Crenezumab had no effect on plaques or clinical progression | **Alzheimer's Prevention Initiative secondary prevention**: Crenezumab had no effect on plaques or clinical progression |
| Plaque Aβ-antibody | **AHEAD A45 secondary prevention:** Ongoing trial of lecanemab in asymptomatic amyloid plaque cohort; clinical progression and plaque removal; **TBD** | **DIAN-TU ART:** Lecanemab in asymptomatic participants from **DIAN-TU Gantenerumab OLE**, amyloid removal in asymptomatic cohort suggests 50% risk reduction in the longest treated group. Study will confirm findings, determine effects of duration of treatment and amyloid removal, and safety of long-term anti-amyloid plaque removal. **Confirmation TBD** |
| | **TRAILBLAZER-ALZ4 secondary prevention:** Ongoing trial of donanemab in asymptomatic amyloid plaque cohort; **TBD** | |
| | **GRADUATE 1 & 2 symptomatic:** Gantenerumab had a moderate effect on plaque removal, non-sig trend clinical benefit | **DIAN-TU Gantenerumab:** In symptomatic participants, non-significant trend for clinical benefit and moderate change in biomarker and amyloid removal |

Concordant completed trials are in green, showing similar outcomes in DIAD and SAD, pending trials are in yellow. TBD: to be determined, studies ongoing.

**Supplemental Table 8. Conversion from abnormal to normal amyloid-plaque status from gantenerumab OLE baseline to the last post-baseline OLE visit in PiB-PET status based on OLE only Gant mITT population.** A higher percentage of gantenerumab prior treated cohort converted from having abnormal levels of amyloid plaques to normal levels compared to placebo or solanezumab previously treated cohorts.

| | | **OLE effect by prior Double-Blind Treatment** | | | |
|---|---|---|---|---|---|
| **Analysis Visit Window Shift** | **Statistic** | **Prior Placebo** | **Prior Solanezumab** | **Prior Gantenerumab** | **Total** |
| Last Post-baseline Value | n | 13 | 21 | 20 | 54 |
| Normal to Normal | n (%) | 1 (8%) | 3 (14%) | 4 (20%) | 8 (15%) |
| Normal to Abnormal | n (%) | 0 | 0 | 0 | 0 |
| Abnormal to Normal | n (%) | 1 (8%) | 2 (10%) | 5 (25%) | 8 (15%) |
| Abnormal to Abnormal | n (%) | 11 (85%) | 16 (76%) | 11 (55%) | 38 (70%) |
| | *p*-value | 0.35 | 0.38 | 0.40 | 0.50 |

**Supplemental Table 9. Mean change of outcomes from OLE baseline (SE) and p-value from MMRM model by year.**

| | **Change from OLE baseline Mean (SE) *p*-value** | | |
|---|---|---|---|
| **Outcome measure** | **Year 1** | **Year 2** | **Year 3** |
| Centiloid | -5.6 (2.5)<br>0.028 | -22.1 (3.1)<br><0.0001 | -29.3 (4.6)<br><0.0001 |
| PiB PET SUVR | -0.12 (0.044)<br>0.012 | -0.47 (0.066)<br><0.0001 | -0.71 (0.088)<br><0.0001 |
| CSF Aβ42/40 CentiMarker | -7.8 (1.88)<br>0.0001 | -29.6 (3.5)<br><0.0001 | -42.0 (5.2)<br><0.0001 |
| CSF Aβ42/40 (%) | 0.67 (0.164)<br>0.0001 | 2.51 (0.305)<br><0.0001 | 3.57 (0.450)<br><0.0001 |
| CSF pTau181/Tau181 CentiMarker | -7.8 (1.4)<br><0.0001 | -17.6 (1.8)<br><0.0001 | -21.0 (2.1)<br><0.0001 |
| CSF pTau181/Tau181 (%) | -2.18 (0.384)<br><0.0001 | -4.84 (0.506)<br><0.0001 | -5.76 (0.595)<br><0.0001 |

**Supplemental Table 10. Mean change in PiB PET SUVR from double-blind baseline by timepoint.**

| | **PiB PET SUVR change from double-blind baseline,<br>Mean (SE)<br>*p*-value** | | | | | | |
|---|---|---|---|---|---|---|---|
| **Double-blind treatment arm** | **Year 1** | **Year 2** | **Year 4** | **OLE baseline** | **OLE Year 1** | **OLE Year 2** | **OLE Year 3** |
| Placebo | 0.14 (0.067)<br>0.041 | 0.19 (0.076)<br>0.018 | 0.44 (0.106)<br>0.0001 | 0.70 (0.150)<br><0.0001 | 0.61 (0.146)<br>0.0001 | 0.19 (0.196)<br>0.34 | -0.07 (0.216)<br>0.74 |
| Solanezumab | | | | 0.60 (0.140)<br><0.0001 | 0.28 (0.136)<br>0.044 | -0.13 (0.175)<br>0.46 | -0.47 (0.191)<br>0.017 |
| Gantenerumab | -0.01 (0.040)<br>0.81 | -0.09 (0.047)<br>0.073 | -0.30 (0.068)<br><0.0001 | -0.19 (0.101)<br>0.059 | -0.07 (0.111)<br>0.55 | -0.38 (0.140)<br>0.0087 | -0.44 (0.162)<br>0.0086 |

**Supplemental Figure 4. Individual CDR-SB by EYO.** Dummy IDs are provided. Key: yellow = DIAN Obs participants (external control), purple = double-blind placebo treated participants, light blue = double-blind gantenerumab treated participants, blue = gantenerumab OLE period data. Double-blind period for solanezumab treated participants are not shown, only their OLE period data are shown.

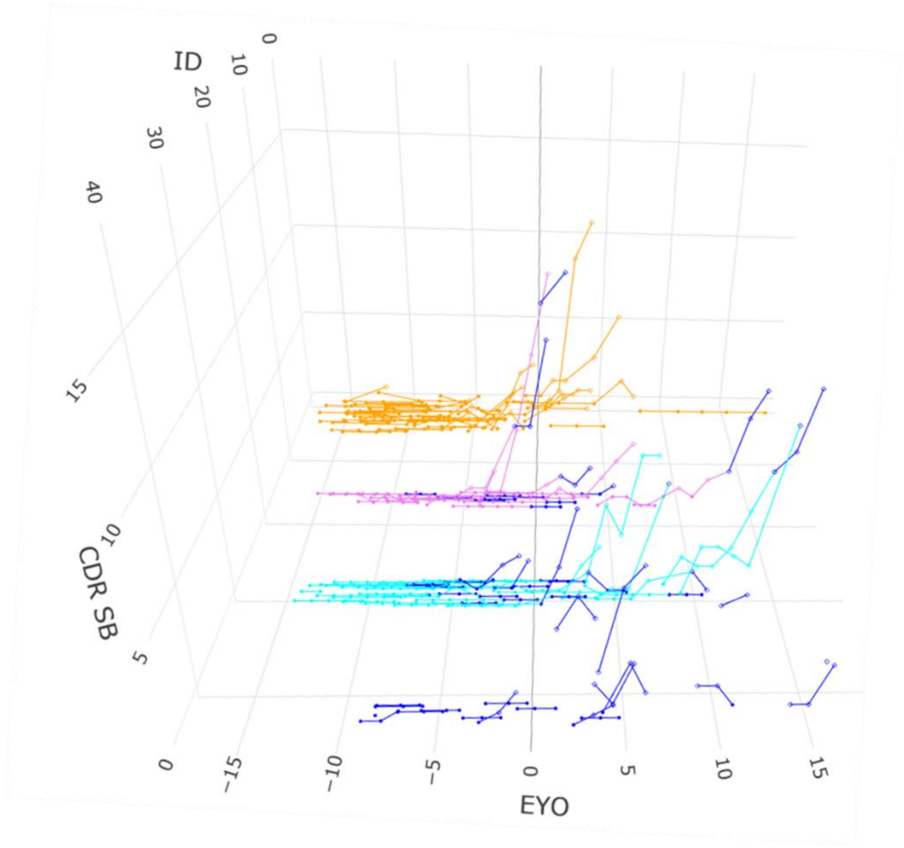

**Supplemental Table 11. Severe treatment emergent adverse events reported.**

| **Treatment emergent adverse event** | **Relationship with Treatment** | **Total** | **Percentage** |
|---|---|---|---|
| Abdominal pain | Not Related | 1 | 1.4% |
| Cholecystitis | Not Related | 1 | 1.4% |
| Colon cancer | Not Related | 1 | 1.4% |
| Costochondritis | Not Related | 1 | 1.4% |
| Metapneumovirus bronchiolitis | Not Related | 1 | 1.4% |
| Musculoskeletal chest pain | Not Related | 1 | 1.4% |
| Psychotic behaviour | Not Related | 1 | 1.4% |
| Sciatica | Not Related | 1 | 1.4% |
| Vomiting | Not Related | 1 | 1.4% |
| Amyloid related imaging abnormality-oedema/effusion | Related | 2 | 2.7% |
| Headache | Related | 1 | 1.4% |

**Supplemental Figure 5. Study populations. A. Participants treated with gantenerumab in either the double-blind or OLE period (Any Gant) were compared to the main set control (internal placebo treated only and external matched controls from DIAN Observational (DIAN Obs) study.** Baseline for this population was defined as double-blind baseline for original gantenerumab treated participants and OLE baseline for original placebo and solanezumab treated participants. B. Participants that were treated with gantenerumab for the longest duration in both the double-blind and OLE period (Longest Gant) and compared to the main set control or the extended control group (dashed box) which are defined as the main set controls plus the double-blind data and OLE baseline values for participants treated with placebo during the double-blind period who entered the OLE. Double-blind baseline was defined as the baseline for this population. C. Participants that were treated with gantenerumab in the OLE period (OLE Only Gant) and only their OLE period data were compared to the main set control. OLE baseline was defined as the baseline for this population.

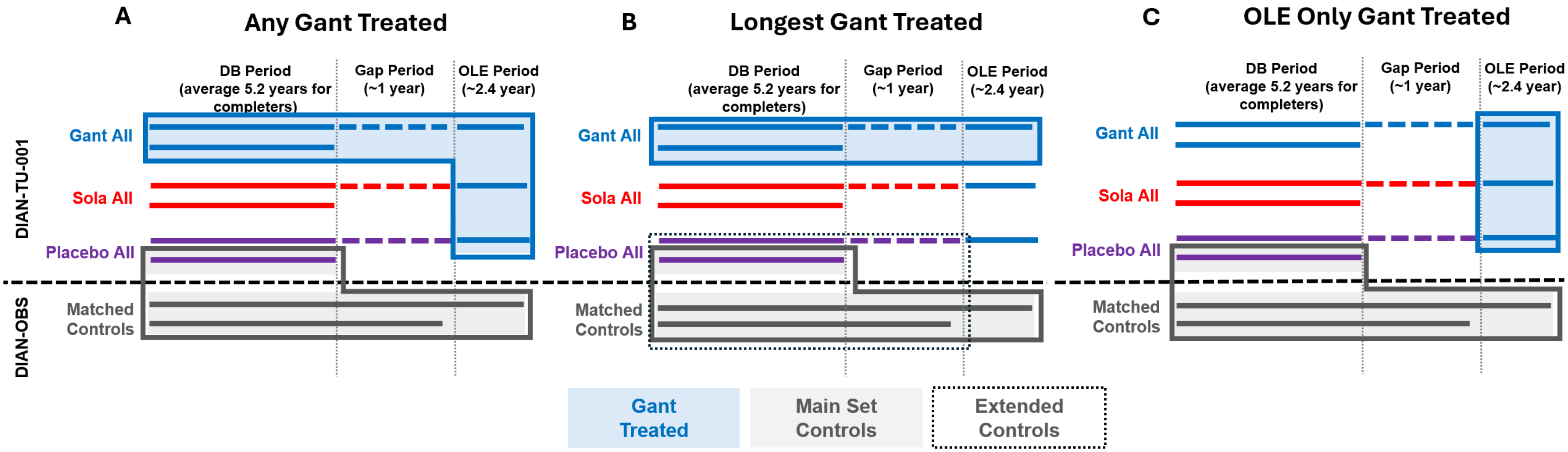

**Supplemental Figure 6. Amyloid levels at the end of the OLE as measured by PiB PET.** Most participants in the primary analysis group (Any Gant) were within normal amyloid range (<25 Centiloids) at the end of the study, although some participants didn't have complete removal of amyloid. In the primary (Any Gant) asymptomatic group (n=52) who were treated with high dose gantenerumab, n=34 (65%) reached Centiloid <25. In all the Any Gant group, n=39 (44%) reached Centiloid <25. Blue dots in the figure represents double-blind baseline asymptomatic longest gantenerumab treated participants.

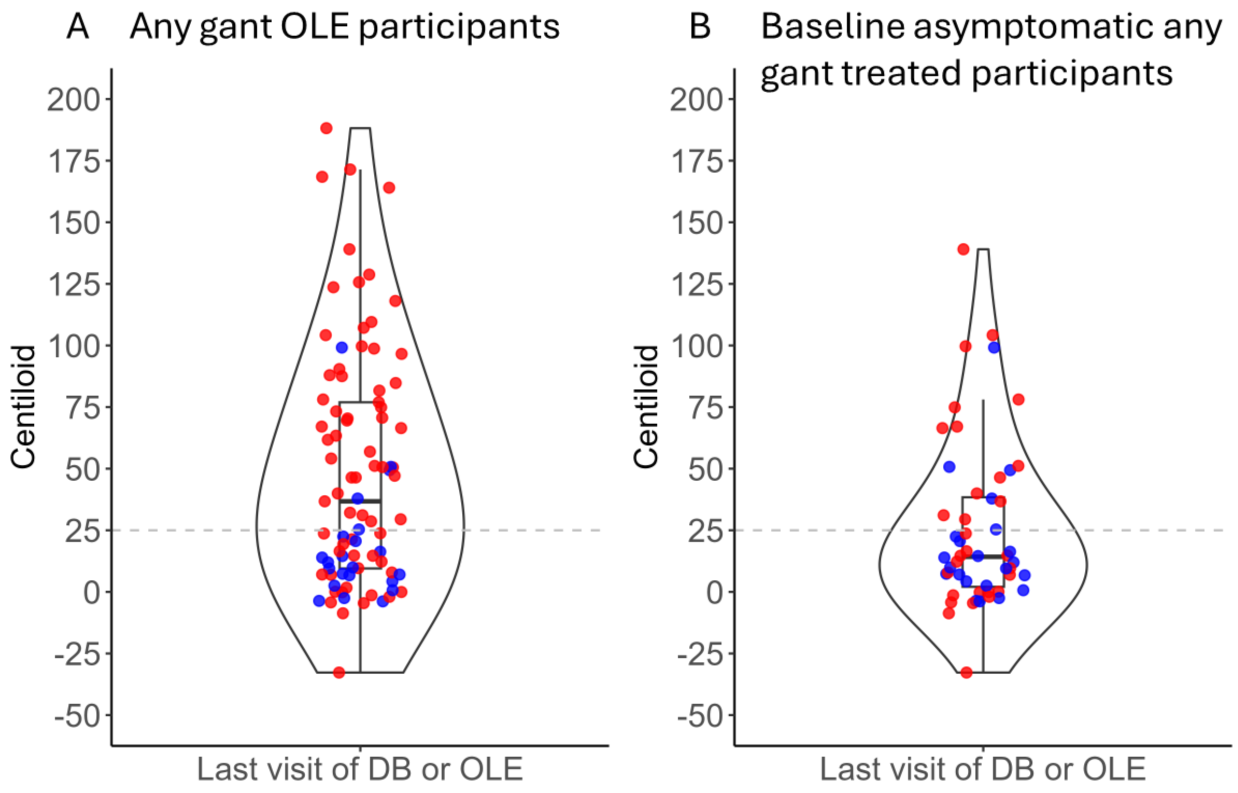

**Supplemental Figure 7. Participant flow to analysis groups. Flow of participants who were asymptomatic at the start of the double-blind (DB) period until the end of the open-label extension (OLE).** Numbers are the sample size (n) in each treatment arm: gantenerumab (Gant), solanezumab (Sola) and placebo. Duration of treatment shown in years. B. Participants who met inclusion criteria and were analysed according to the pre-specified primary analyses are shown. Asymptomatic mutation carriers at the DIAN-TU-001 randomized, double blind (DB) period baseline were off treatment for approximately 1 year before starting the open label extension study when all were assigned to gantenerumab. The two cohorts included those who were treated the longest time period with gantenerumab (n=22 of 31 starting) and those treated the shortest time (n=23 of 54 starting) with gantenerumab dosing duration average of 8.4 versus 2.7 years respectively.

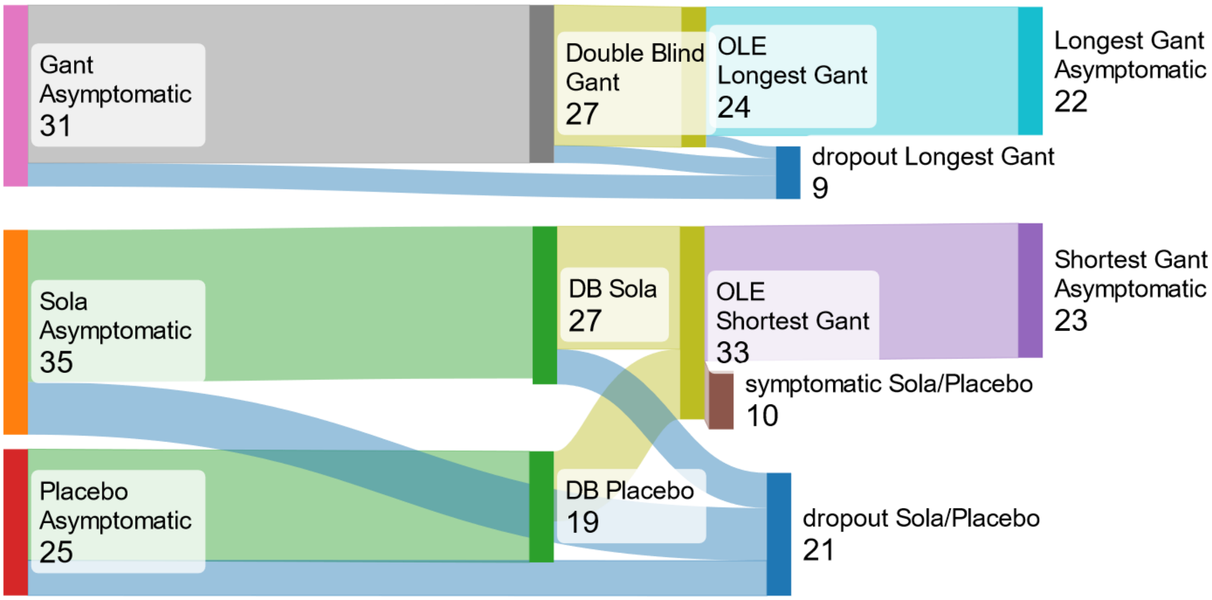
